# Multi-ancestry GWAS of severe pregnancy nausea and vomiting identifies risk loci associated with appetite, insulin signaling, and brain plasticity

**DOI:** 10.1101/2024.11.19.24317559

**Authors:** Marlena Fejzo, Xinran Wang, Julia Zöllner, Natàlia Pujol-Gualdo, Triin Laisk, Estonian Biobank Research Team, Sarah Finer, David A. van Heel, Genes & Health Research Team, Ben Brumpton, Laxmi Bhatta, Kristian Hveem, Elizabeth A. Jasper, Digna R. Velez Edwards, Jacklyn N. Hellwege, Todd Edwards, Gail P. Jarvik, Yuan Luo, Atlas Khan, Kimber MacGibbon, Yuan Gao, Gaoxiang Ge, Inna Averbukh, Erin Soon, Michael Angelo, Per Magnus, Stefan Johansson, Pål R. Njølstad, Marc Vaudel, Chang Shu, Nicholas Mancuso

**Author notes:** These authors are co-corresponding authors (Marlena Fejzo, Marc Vaudel, Chang Shu, Nicholas Mancuso).

## Abstract

While most pregnancies are affected by nausea and vomiting, hyperemesis gravidarum (HG) is at the severe end of the clinical spectrum and is associated with dehydration, undernutrition, and adverse maternal, fetal, and child outcomes. Herein we performed a multi-ancestry genome-wide association study (GWAS) of severe nausea and vomiting of pregnancy of 10,974 cases and 461,461 controls across European, Asian, African, and Latino ancestries. We identified ten significantly associated loci, of which six were novel (*SLITRK1*, *SYN3*, *IGSF11*, *FSHB*, *TCF7L2*, and *CDH9),* and confirmed previous genome-wide significant associations with risk genes *GDF15*, *IGFBP7*, *PGR*, and *GFRAL*. In a spatiotemporal analysis of placental development, *GDF15* and *TCF7L2* were expressed primarily in extra villous trophoblast, and using a weighted linear model of maternal, paternal, and fetal effects, we confirmed opposing effects for *GDF15* between maternal and fetal genotype. Conversely, *IGFBP7* and *PGR* were primarily expressed in developing maternal spiral arteries during placentation, with effects limited to the maternal genome. Risk loci were found to be under significant evolutionary selection, with the strongest effects on nausea and vomiting mid-pregnancy. Selected loci were associated with abnormal pregnancy weight gain, pregnancy duration, birth weight, head circumference, and pre-eclampsia. Potential roles for candidate genes in appetite, insulin signaling, and brain plasticity provide new pathways to explore etiological mechanisms and novel therapeutic avenues.

## Introduction

Most pregnancies are affected by nausea and vomiting (NVP), but in 0.3-10.8% of pregnancies the symptoms can be severe enough to cause maternal weight loss and adverse maternal and fetal outcomes.^1,2^ The most severe form, Hyperemesis Gravidarum (HG), is the leading cause of hospitalization in the first trimester and the second leading cause, after preterm labor, of pregnancy hospitalization overall.^3^ Current treatments for HG are frequently ineffective in improving patient symptoms, thereby increasing risk of pregnancy termination, suicidal ideation, postpartum depression and numerous other maternal and offspring comorbidities.^4,5,6,7,8,9^ Therefore, understanding of HG etiology is critical to begin to address the negative impact severe NVP has on maternal and child health.

While the historical hypothesis centered around the pregnancy hormone hCG, recent large-scale genetic studies have implicated Growth Differentiation Factor-15 gene (*GDF15*), a hormone associated with nausea and vomiting.^10,11^ *GDF15* was identified as the greatest genetic risk-factor for HG in both a genome-wide and an exome-wide association study, and a rare mutation in *GDF15* was associated with >10-fold increased risk for HG.^10,11^ Other significant genetic associations that have been replicated include the gene coding for the brainstem-restricted receptor for GDF15, GDNF family receptor alpha-like (*GFRAL)*, and the placental proteins: insulin-like growth factor binding protein 7 (*IGFBP7*) and the progesterone receptor (*PGR*).^10^ Despite these significant advances, further work in larger and more diverse populations is necessary to determine the generalizability of these findings and identify additional genetic associations and potential therapeutic targets.

Here, we performed a multi-ancestry GWAS of 10,974 HG/excessive vomiting in pregnancy cases and 461,461 controls across European, Asian, African, and Latino ancestries from 9 contributing studies. GWAS confirmed previous genome-wide significant associations with *GDF15*, *GFRAL*, *IGFBP7*, and *PGR*, and identified six additional loci (*SLITRK1*, *SYN3*, *IGSF11*, *FSHB*, *TCF7L2*, and *CDH9)* significantly associated with HG risk. We analyzed the risk loci with respect to maternal and fetal/paternal contributions, other phenotypic associations, anthropometric measures and pregnancy outcomes, temporal association with nausea and vomiting throughout gestation, and spatiotemporal expression in the developing placenta. Overall, our work provides additional insight into the genetic etiology of HG risk.

## Results

### Multi-ancestry HG GWAS meta-analysis identifies novel risk regions

Our discovery meta-analysis of GWAS on HG encompassed nine independent studies: three cohorts from the United States (23andMe, Inc.,^13^ Fejzo,^11^ and eMERGE^13^), one from Estonia (Estonian Biobank),^14^ two from the United Kingdom (UK Biobank,^15^ and Genes & Health^16^), one from Finland (FinnGen Biobank, Release 8^17^), and two from Norway (HUNT^18^ and MoBa^19^). In total, we analyzed 10,974 cases and 461,461 controls across European (N=456,817), Asian (N=13,159), African (N=1,277), and Latino (N=75) ancestries (see **Table 1**). Broadly, cases were defined as patients diagnosed with HG or excessive vomiting during pregnancy, and controls were defined as females with no NVP diagnoses (see **Table S1**). For each contributing study we performed rigorous QC on genotyped and imputed variants, summary statistics, and outlier substudies (i.e., studies contributing to individual biobank results; see **Methods**).

**Table 1.** Source population. Ancestries include European (EUR), African (AFR), Asian (East and South Asian combined), Admixed American.

| SOURCE | CASE (N) | CONTROL (N) | EUR | AFR | ASIAN | AMR | UNK* |
| --- | --- | --- | --- | --- | --- | --- | --- |
| 23andMe, Inc. | 1,306 | 15,756 | 15,892 | 0 | 0 | 0 | 0 |
| Fejzo | 926 | 660 | 1506 | 43 | 25 | 75 | 24 |
| FinnGen R8 | 2,092 | 163,702 | 165,794 | 0 | 0 | 0 | 0 |
| Estonian Biobank | 3,536 | 32,113 | 35,649 | 0 | 0 | 0 | 0 |
| UK Biobank | 220 | 180,917 | 181,137 | 0 | 0 | 0 | 0 |
| Genes & Health | 1,544 | 11,128 | 0 | 0 | 12,672 | 0 | 0 |
| HUNT | 269 | 36,460 | 36,729 | 0 | 0 | 0 | 0 |
| eMERGE | 504 | 6364 | 5,172 | 1,234 | 462 | 0 | 0 |
| MoBa | 577 | 14,361 | 14,938 | 0 | 0 | 0 | 0 |
| **TOTAL** | **10,974** | **461,461** | **456,817** | **1,277** | **13,159** | **75** | **24** |

\*Unknown (Other) ancestry reported using Regeneron Genetics Pipeline determined as described previously (11).

We first performed genome-wide association scans for the six contributing studies with individual-level data (see **Table S1**) using logistic mixed-effect models. Next, we performed a fixed-effects meta-analysis using summary statistics from autosomes of all nine cohorts and sex chromosomes where available resulting in total of 43,961,852 examined variants (see **Methods**). Overall, we identified ten independent genome-wide significant loci (*p* < 5×10^-8^; see **Figure 1**). Of these 10 loci, six were novel (defined as at least 1 Mb distance from any other genome-wide significant association; see **Figure 1**, **Table 2**, **Figures S1-S2**. The genomic inflation factor λ_GC_was 1.16 (**Figure S3**). No variant demonstrated genome-wide significance for heterogeneity across studies (*p* < 5×10^-8^; see **Table 2, Methods**).

**Figure 1.**
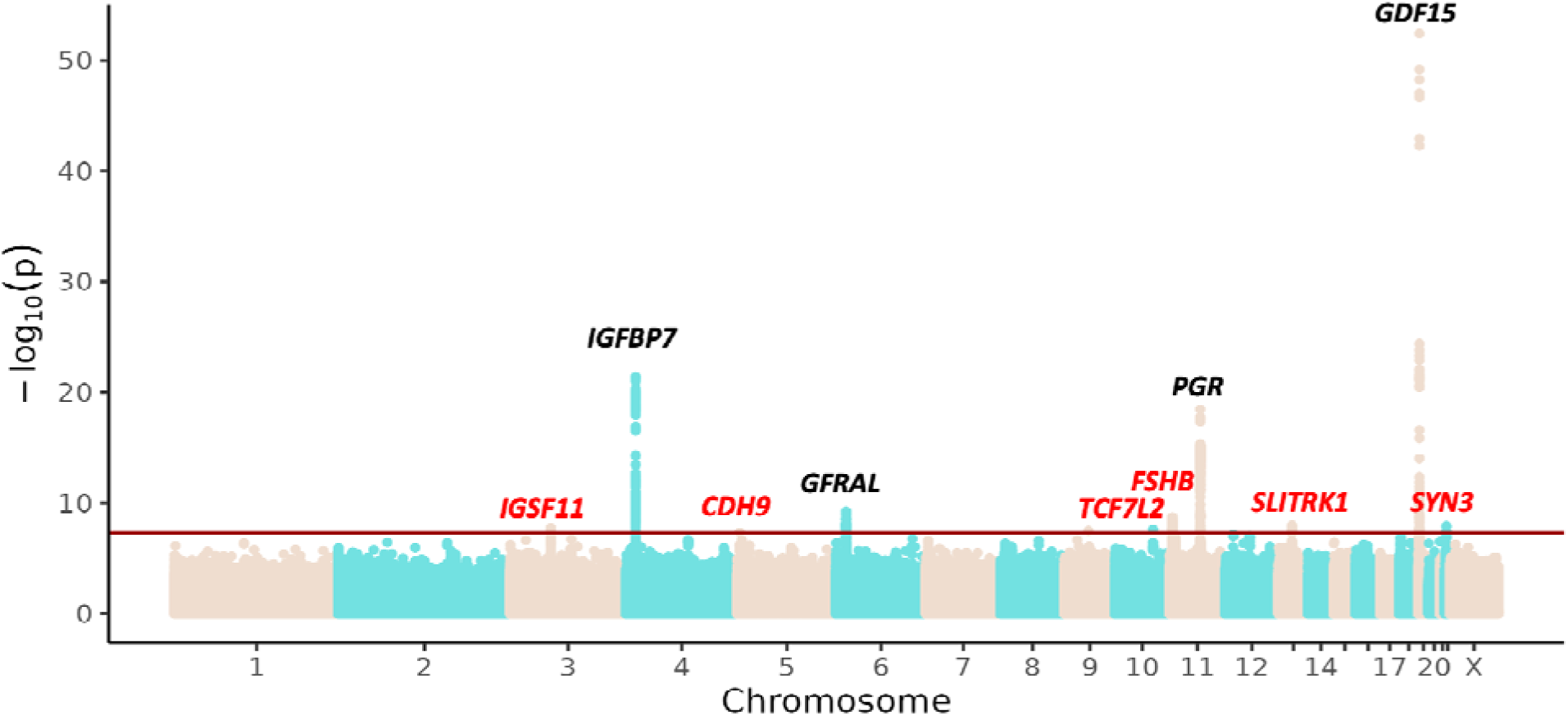
Manhattan plot for HG multi-ancestry GWAS. The plot shows -log10 transformed *p*-value against genomic positions. The red line represents genome-wide significant threshold *p* < 5×10^−8^. The significant SNPs are annotated with the nearest candidate gene. In the annotation, red indicates novel SNPs and black indicates SNPs replicated in previous studies.

**Table 2.**
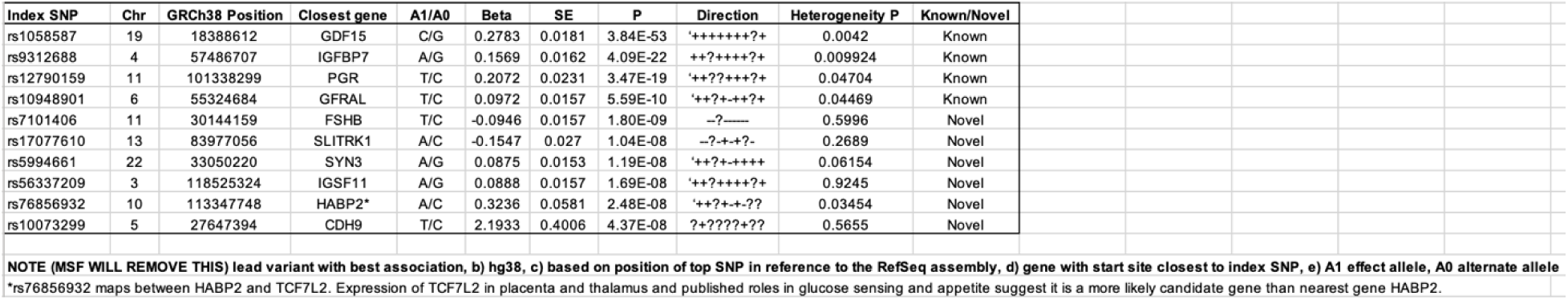
Lead SNPs for multiethnic meta-analysis of 9 GWASes. Direction from left to right corresponds to cohorts finngen, 23andMe, Fejzo Estonian.

Next, we performed statistical fine-mapping for each significant 1Mb region to identify independent signals and putative causal variants. Briefly, we used susieR^20^ to perform statistical fine-mapping of meta-analysis summary statistics at identified risk regions, using estimates of linkage disequilibrium (LD) from reference panels. Before fine-mapping, we further pruned summary statistics to sites having MAF ≥ 0.01 and non-ambiguous alleles (e.g., A/T, C/G) with exceptions for lead variants (see **Methods**). Across 9/10 risk regions, fine-mapping identified 14 95%-credible sets, with 7/9 fine-mapped regions containing a single credible set, suggesting mild allelic heterogeneity underlying HG risk regions. Credible sets contained 20 SNPs on average consistent with pervasive LD patterns at GWAS risk regions, however 3/14 credible sets were comprised of an individual SNP (see **Table S2, Table S3, Figure S2**). Fine-mapping at the risk region around *CDH9* (rs10073299, chr5:27647394) failed to produce a credible set, due to GWAS signals being pruned due to MAF thresholds.

### Joint Maternal and Fetal Analysis confirms Opposing Contributions for GDF15

Next, we investigated the relative contribution of maternal and fetal genetics to the etiology of HG in the Norwegian Mother, Father and Child cohort study (MoBa), a pregnancy-based prospective cohort representing more than 100,000 pregnancies. Briefly, we leveraged a recent approach^21^ that estimates the contribution of parental/fetal effects by analyzing GWASs conducted against parents and children separately (see Methods).^21^ Among the ten genome-wide significant loci identified in the meta-analysis, nine were genotyped with high quality in MoBa and selected for downstream analysis.

For the SNPs with nearest genes *IGFBP7*, *FSHB*, *IGSF11*, *PGR*, and *TCF7L2-HABP2*, the effect was primarily carried by the maternal genome. At our current sample size, the confidence interval of the fetal effect was crossing the null for all these loci (**Figure 2**). We found opposite allele effects for rs1058587 in *GDF15* on HG risk between mother and fetus, in line with the hypothesis that GDF15-increasing alleles increase risk through expression from the fetal genome in early pregnancy but provide protection through pre-pregnancy exposure in the maternal genome.^22^ Interestingly, the maternal effect was stronger than the fetal effect, suggesting that the maternal pre-pregnancy habituation to GDF15 may prevail over fetal overexpression during pregnancy

**Figure 2.**
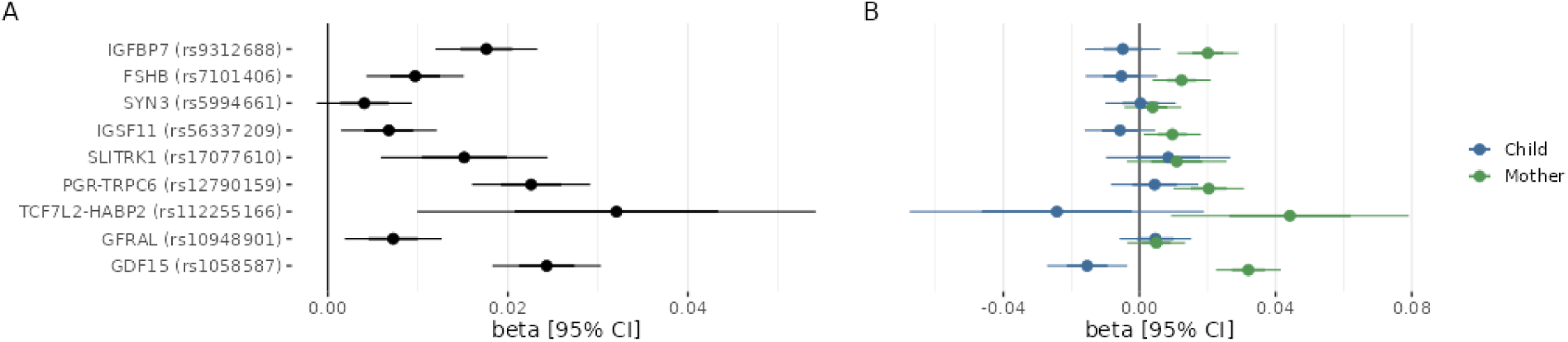
Effect size estimates for the lead SNP at each locus in the MoBa cohort. Forest plot of the effect size estimates when encoding no NVP, NVP, and HG as 0, 1, and 2, respectively, for (A) maternal effect estimates as obtained from the GWAS against the maternal genome and (B) conditioned maternal and fetal effects obtained after WLM-adjustment of effect estimates in GWASes against the fetal, maternal, and paternal genomes. For the sake of readability, paternal effect sizes are not represented. Points represent effect size estimates and error bars represent 95% confidence intervals of the estimate.

### HG risk is enriched for regions under constraint

To characterize the genetic architecture of HG, we performed partitioned LD score regression using the meta-analysis summary statistics (see Methods). First, LDSC estimated an intercept of 1.0281 (SE: 0.0086), suggesting results may be affected in part by population stratification.

Second, LDSC estimated significant heritability on a liability scale 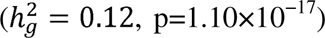 when provided 2.3% HG case prevalence in our study samples and 2% population prevalences. To account for varying reports of population prevalence, we re-analyzed our results using differing values and found broadly consistent estimates of 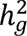 on the liability scale (**see Table S4**). When partitioning 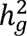 across various functional and evolutionary categories, we found significant associations between measures of gene constraint with HG heritability (*p* <0.0005; see **Figure 3**). For example, GERP^23^ scores, suggesting highly disproportionate conservation and providing evidence that HG risk loci are under evolutionary constraint.

**Figure 3.**
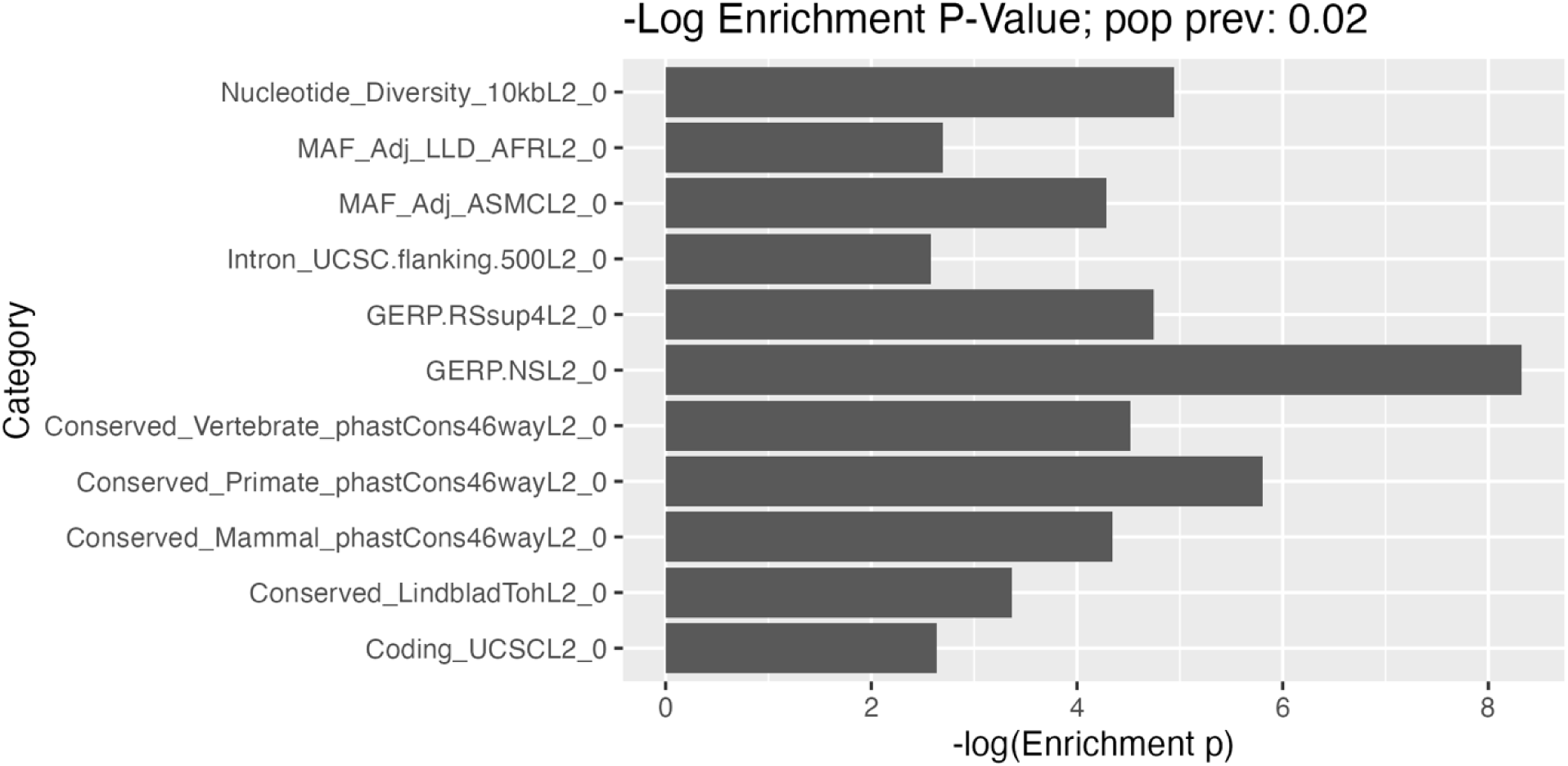
Enrichment analysis suggests evolutionary constraint shapes genetic architecture of HG.

### HG risk genes linked to female and pregnancy characteristics and outcomes

To understand the broader phenotypic consequences of HG risk variants identified in this study, we performed a PheWAS for each of the 10 genome-wide significant SNPs in our meta-analysis using the OpenTargets platform (see Methods). After adjusting for multiple testing (*p* < 1.48×10^−6^), 3/10 variants exhibited associations with additional phenotypes, including traits involved in GDF15 measurements (rs1058587), body size (e.g., fat percentage; rs1058587), female reproductive system (e.g., such as length of menstrual cycle, endometriosis, and ovarian cysts; rs7101406), and chronotype (e.g., getting up in the morning; rs10948901; **Figure 4**).

**Figure 4.**
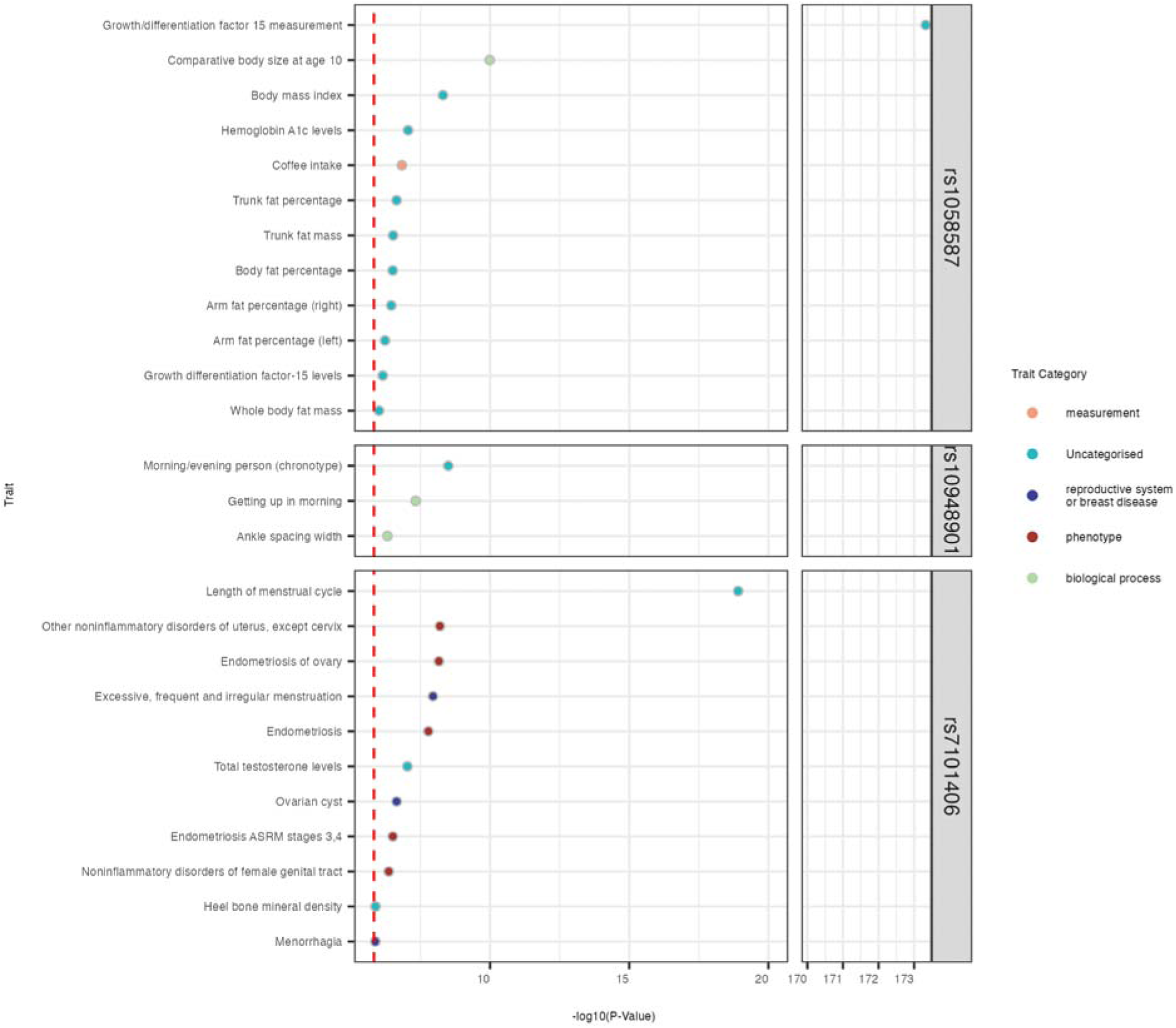
Phenome-wide association study (PheWAS) on lead HG variants. Phenotypes were identified for 3 / 10 variants (*p* < 1.48×10^−6^ after adjusting for multiple testing; red dashed line).

Next, we sought to explore the phenotypic consequences of these variants focusing on maternal characteristics (maternal weight/weight gain/BMI at beginning of pregnancy, at 15 weeks’ gestation, and at term) in HG cases and controls in the MoBa cohort (see **Table 3 and Supplementary Table S5)**. Overall, we identified associations between risk loci and maternal weight and BMI at the beginning of pregnancy (*GDF15*, *SLITRK1*), at 15 weeks (*GDF15*), and at the end of pregnancy (*GDF15*, *PGR*). Risk loci were also associated with maternal weight gain at 15 weeks (most strongly associated with *GDF15*, but also associated with *IGFBP7* and *PGR*) and at the end of pregnancy (most strongly associated with *PGR-TRPC6*, but also associated with *IGFBP7* and *GDF15*). Overall, the PheWAS and MoBa studies suggest risk loci may contribute to maternal weight and/or body size prior to and throughout gestation.

**Table 3.** Individual SNP associations with pregnancy outcomes and maternal health as reported by the mother or recorded in the Norwegian Medical Birth Registry (NMBR).

| SNP | Allele | Phenotype | Source | Beta | SE | N | Locus | P-value |
| --- | --- | --- | --- | --- | --- | --- | --- | --- |
| rs12790159 | C | Preeclampsia or eclampsia | NMBR | 0.154 | 0.048 | 44436 | PGR/TRPC6 | 1.30E-03 |
| rs1058587 | G | Preeclampsia labeled as serious | NMBR | 0.23 | 0.085 | 44197 | GDF15 | 6.70E-03 |
| rs12790159 | C | Pregnancy duration in days | NMBR | -0.388 | 0.093 | 44261 | PGR/TRPC6 | 2.90E-05 |
| rs12790159 | C | Birthweight adjusted for sex and pregnancy duration | NMBR | -0.014 | 0.004 | 44401 | PGR/TRPC6 | 9.80E-04 |
| rs56337209 | G | Head circumference adjusted for sex and pregnancy duration | Self-reported | 0.038 | 0.01 | 43462 | IGSF11 | 2.00E-04 |
| rs12790159 | C | Head circumference adjusted for sex and pregnancy duration | Self-reported | -0.034 | 0.013 | 43722 | PGR/TRPC6 | 7.00E-03 |
| rs17077610 | A | Maternal weight in kg at the beginning of pregnancy | Self-reported | -0.412 | 0.154 | 41759 | SLITRK1 | 7.60E-03 |
| rs1058587 | G | Maternal weight in kg at the beginning of pregnancy | Self-reported | 0.286 | 0.101 | 41472 | GDF15 | 4.50E-03 |
| rs1058587 | G | Maternal weight in kg at week 15 of gestation | Self-reported | 0.461 | 0.1 | 40470 | GDF15 | 4.30E-06 |
| rs12790159 | C | Maternal weight in kg at the end of pregnancy | Self-reported | 0.394 | 0.122 | 36705 | PGR/TRPC6 | 1.20E-03 |
| rs1058587 | G | Maternal weight in kg at the end of pregnancy | Self-reported | 0.541 | 0.112 | 36497 | GDF15 | 1.20E-06 |
| rs17077610 | A | Maternal BMI in kg/m2 at the beginning of pregnancy | Self-reported | -0.134 | 0.052 | 41640 | SLITRK1 | 9.80E-03 |
| rs1058587 | G | Maternal BMI in kg/m2 at the beginning of pregnancy | Self-reported | 0.096 | 0.034 | 41357 | GDF15 | 4.40E-03 |
| rs1058587 | G | Maternal BMI in kg/m2 at week 15 of pregnancy | Self-reported | 0.153 | 0.033 | 40371 | GDF15 | 4.50E-06 |
| rs12790159 | C | Maternal BMI in kg/m2 at the end of pregnancy | Self-reported | 0.151 | 0.041 | 36013 | PGR/TRPC6 | 1.90E-04 |
| rs1058587 | G | Maternal BMI in kg/m2 at the end of pregnancy | Self-reported | 0.196 | 0.037 | 35814 | GDF15 | 1.40E-07 |
| rs9312688 | G | Maternal weight gain in kg at week 15 of gestation | Self-reported | 0.078 | 0.024 | 40497 | IGFBP7 | 1.10E-03 |
| rs12790159 | C | Maternal weight gain in kg at week 15 of gestation | Self-reported | 0.133 | 0.028 | 40507 | PGR/TRPC6 | 1.60E-06 |
| rs1058587 | G | Maternal weight gain in kg at week 15 of gestation | Self-reported | 0.183 | 0.025 | 40298 | GDF15 | 5.80E-13 |
| rs9312688 | G | Maternal weight gain in kg at the end of pregnancy | Self-reported | 0.167 | 0.049 | 35533 | IGFBP7 | 7.40E-04 |
| rs12790159 | C | Maternal weight gain in kg at the end of pregnancy | Self-reported | 0.403 | 0.057 | 35556 | PGR/TRPC6 | 2.10E-12 |
| rs1058587 | G | Maternal weight gain in kg at the end of pregnancy | Self-reported | 0.222 | 0.053 | 35359 | GDF15 | 2.40E-05 |

Since HG has been associated with adverse pregnancy and offspring outcomes such as increased risk for preeclampsia and smaller brains in offspring,^2,24^ we explored the association between the HG risk genes and pregnancy characteristics and outcomes (preeclampsia, gestational diabetes, pregnancy duration, and placental weight) and offspring characteristics and outcomes (perinatal death, birth weight, head circumference). According to the Medical Birth Registry of Norway (MBRN), in the MoBa cohort there was a 5-fold increased risk of early preeclampsia in HG cases that went to term compared to those with no NVP, and a 3-fold increased risk of early preeclampsia for HG cases that were born less than 37 weeks’ gestation, but the low number of cases did not allow reaching statistical significance. Among the risk loci identified in the meta-analysis included in MoBa, PGR-TRPC6 was moderately associated with preeclampsia (p = 1.3×10^−3^) and GDF15 with preeclampsia labeled as ‘serious’ in the MBRN (p = 6.7×10^−3^). Phenotypically, children born to mothers with HG were lighter at birth, had a heavier placenta for term birth, but not for preterm birth (**Supplementary Table S5**). A low or high placental growth can be a risk factor for early- and late-onset preeclampsia, respectively.^25^ Placental growth imbalance therefore makes a possible link between HG and preeclampsia, yet none of the variants found in this study was associated with placental weight in MoBa. However, the PGR-TRPC6 risk variant was associated with pregnancy duration (p = 2.9×10^−5^) and birthweight (p = 9.8×10^−4^) and both PGR-TRPC6 (p = 7.0×10^−3^) and IGSF11 (p = 2.0×10^−4^) were associated with offspring head circumference (**Table 3**). No associations were identified between HG risk genes and perinatal death nor gestational diabetes.

### Selected loci have greatest effects on NVP symptoms at different gestational ages, with overall greatest effects on NVP at 13-20 weeks’ gestation

Despite the majority of NVP symptoms resolving by the end of the first trimester, HG lasts until term in 22% of cases.^26^ To determine whether risk loci have an effect on duration of symptoms, we estimated effects on NVP cross sectionally during pregnancy starting prior to gestational age four weeks up until ≥29 weeks in the MoBa cohort (**Figure 5**). We found SNPs at loci *GDF15*, *PGR-TRPC6*, and *IGFBP7* exhibited their greatest effects between gestational weeks 13-20 with effects of *GDF15* and *PGR-TRPC6* extend all the way to term. Interestingly, the effect of *GFRAL* exhibited only during the second trimester of pregnancy, which is consistent with GFRAL desensitization explaining the absence of nausea and vomiting in the second half of pregnancy in the majority of pregnancies despite increasing GDF15 levels.^22^ Estimated effects for the remaining loci varied. For example, *SLITRK1* and *IGSF11* demonstrated strongest effects in the first trimester, while *TCF7L2-HABP2* and *SYN3* affected HG risk later in pregnancy. Overall, we find evidence for a temporal component to the genetic architecture of HG risk.

**Figure 5.**
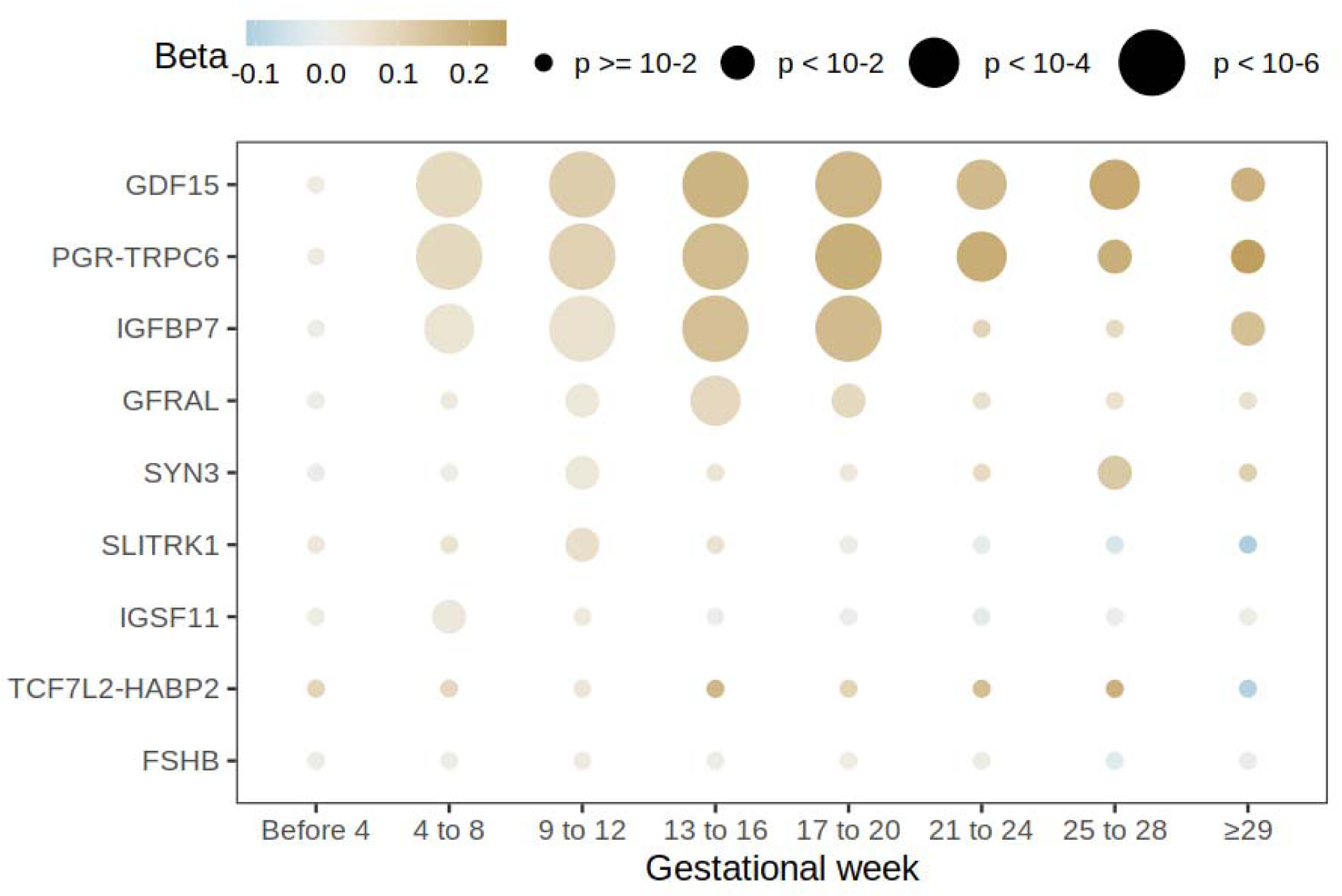
Cross sectional analysis in MoBa. Maternal effect size estimates of the lead SNPs obtained in GWASes of NVP reported by mothers stratified by gestational week. The size of the point represents the significance of the association and its color the effect size estimate.

### Spatiotemporal candidate gene expression during early placental development

Several of the genes associated with HG have previously been reported to be expressed during early placental development.^23^ Therefore, using a spatial transcriptomics approach, we analyzed the spatiotemporal expression of genes proximal to HG risk loci in fetal tissue and maternal tissue of the decidua during trophoblast differentiation, invasion, and spiral artery remodeling. The highest levels of normalized expression in extra-villous trophoblasts of the fetal placenta and/or the spiral arteries of the developing maternal decidua were found for *GDF15*, *IGFBP7*, *TCF7L2*, and *PGR* (**Figure 6**). Consistent with our previous publication showing the majority of circulating GDF15 comes from the fetus during pregnancy,^24^ GDF15 was not detected in the maternal spiral arteries and decidua. In the developing fetal placenta, the highest level of expression of *GDF15* was detected in the anchoring Extravillous trophoblasts (EVTA), with expression detected in Floating Villi, Villous cytotrophoblasts, and Interstitial Extravillous trophoblasts (EVTI), but *GDF15* expression decreased significantly in Endovascular Extravillous trophoblasts (EVTE) once EVT are inside the arteries. *TCF7L2* was expressed at high levels in EVTA with expression peaking in EVTI during decidual invasion, and lower levels detected in EVTE. Albeit at relatively lower levels, *TCF7L2* expression was detected in maternal decidua with low levels in spiral arteries. Conversely, expression of *IGFBP7* and *PGR* was limited to the maternal side with expression in spiral arteries declining with remodeling. Spatial or temporal trends during placentation were not detected for the remaining risk loci.

**Figure 6.**
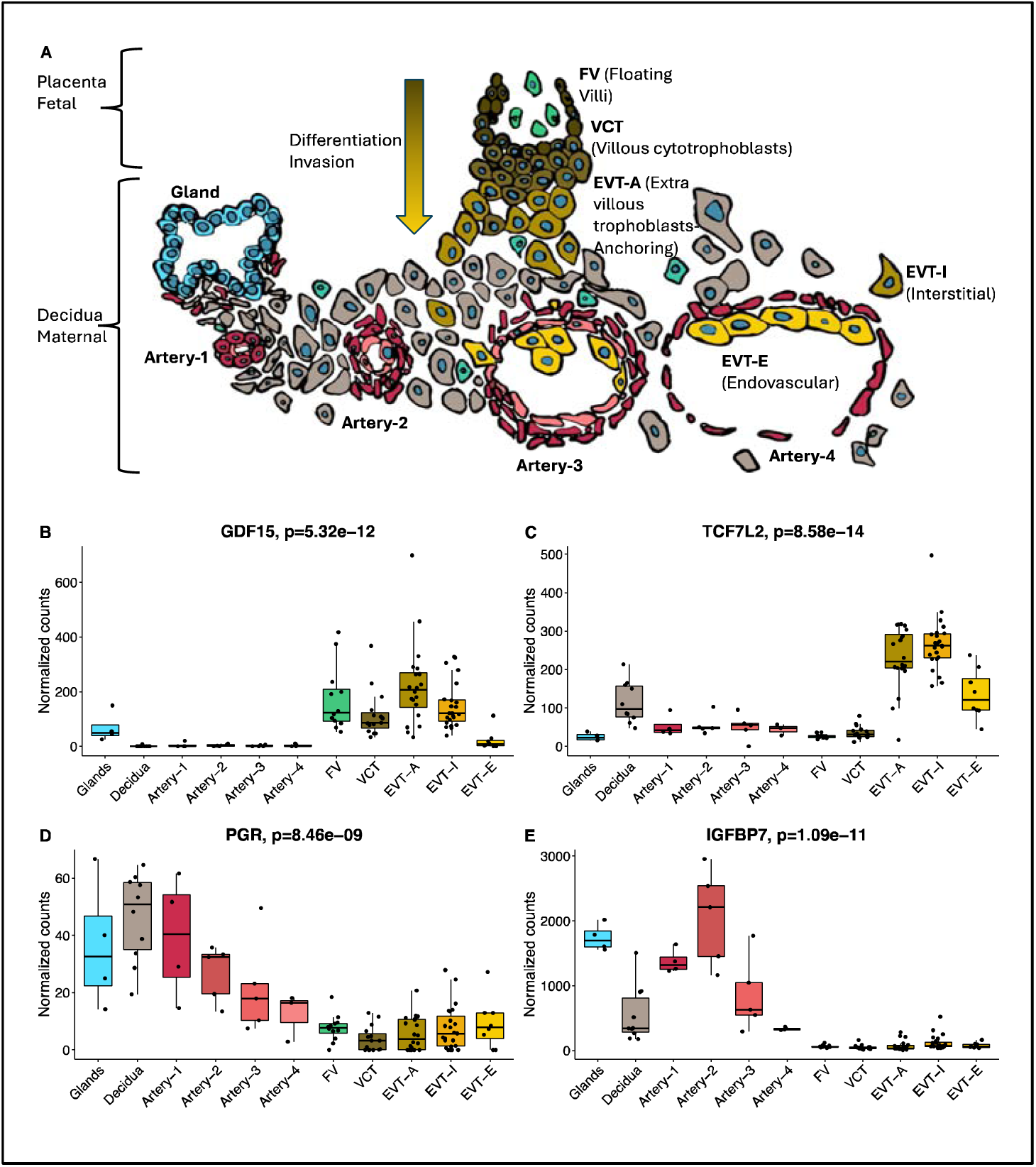
Spatiotemporal candidate gene expression during early placental development. A) Model of fetal and maternal tissue during trophoblast differentiation, invasion, and spiral artery remodeling. Regions of interest (ROI) were selected corresponding to one of the following categories: artery (N=17), decidua (N=10), floating villi (N=12), VCT (N=15), interstitial EVT (N=23), anchoring EVT (N=20), endovascular EVT (N=8), endometrial glands (N=4) from 30 patients between 6-20 weeks’ gestation. Normalized expression (individual gene counts divided by counts for all genes) for each ROI is shown for B) GDF15, C) TCF7L2, D) PGR, and E) IGFBP7.

### Novel candidate genes are differentially expressed in brain

Six novel loci with candidate genes expressed in the brain were identified in this study by searching protein and mRNA expression datasets shown in **Supplementary Table S6**. Among them, nearest genes *SYN3* (Synapsin-3), *SLITRK1* (SLIT And NTRK Like Family Member 1), and *IGSF11* (Immunoglobulin Superfamily Member 11) all have enriched (≥4-fold higher expression in brain compared to any other tissue analyzed) or enhanced (moderately elevated) expression in brain. Of note, novel genes *TCF7L2* (transcription factor 7-like 2, also known as *TCF4*) and *FSHB* (follicle stimulating hormone subunit beta) are specifically enriched in the thalamus (brain region involved in feeding behavior) and the pituitary gland (brain region involved in reproduction), respectively.

### Functional analysis of the most significant risk variant, rs1058587

Among the four loci identified in this study that have been replicated in previous GWAS studies, rs1058587 (also known as the H6D polymorphism because of the variation at position 6 of the mature Growth and Differentiation Factor 15 protein, GDF15) was the most significant risk locus for HG. An SNP at the *GDF15* locus was the lead signal in every individual cohort-level GWAS/EWAS included in this study that had a statistically significant association (23andMe, Fejzo, Estonian Biobank, Genes & Health, and FinnGen Biobank), suggesting its generalizability in predisposing to HG. The rs1058587 variant (H in H6D) is associated with lower circulating levels of the hormone.^24^ We found a strong association between rs1058587 and GDF15 measurement (P=4.73×10^-174^) (**Figure 3**), but how this variant results in the profound alteration in circulating GDF15 levels is unknown. The H6D maps close to the cleavage site in the pro-peptide. Therefore, we and previous researchers hypothesized that the H-variant may result in reduced cleavage to mature protein as a mechanism to explain the very significant association with lower circulating levels for this variant.^27^ To test this, we compared the ratio of secreted processed pro-peptide to precursor for both variants in an in vitro assay. No significant difference was identified suggesting an alternate mechanism or linked causal SNP to explain the difference in GDF15 levels between the two variants (**Supplementary Figure S2**).

## Discussion

This GWAS meta-analysis of HG validated four known associations and identified six novel associations that map within or in close proximity to candidate genes *FSHB, SLITRK1, SYN3, IGSF11, TCF7L2,* and *CDH9.* Since historically the pregnancy hormones hCG and estrogen have both been hypothesized to be causal candidates for HG,^26^ it is important to emphasize that the meta-GWAS did not identify any association with the genes encoding these hormones nor their receptors, but rather, identified a novel association with the follicle-stimulating hormone subunit beta (FSHB), and validated the strong association with the nausea and vomiting hormone (GDF15), its receptor (GFRAL), and the hormone receptor for progesterone (PGR). Future work linking hormones to HG should focus on these hormones, rather than continuing to expend limited resources on studies of hCG and estrogen given the lack of genetic evidence to support a connection. Of note, FSH is expressed in the developing placenta with high expression in maternal decidua at eight and ten weeks’ gestation and almost 10-fold higher expression in term myometrium compared to non-pregnant myometrium.^28^ However, low FSH levels are a risk factor for chemotherapy-induced nausea and vomiting, suggesting there may be an inverse relationship between FSH and HG.^29^ Another possible explanation for the association between FSHB and HG may be indirectly through increasing the risk of multiple gestation since the greatest risk locus for dizygotic twinning is in *FSHB* (rs11031005), which is in linkage disequilibrium (r2=0.2, D’=0.88) with the HG *FSHB* locus (rs7101406), and multiple gestation is an independent risk factor for HG.^30,31^

Among the novel loci identified in this study are three that map within or near brain-enriched genes (*SYN3*, *SLITRK1*, *IGSF11).* These genes did not show enrichment for placental expression and thus we hypothesize these neuronal synapse genes may play a role in downstream signaling of appetite, nausea and vomiting in the brain.^32–34^ A role in appetite is indirectly supported by our finding in this study that *SLITRK1* is associated with pre-pregnancy maternal weight and BMI. Additionally, *SYN3*, *SLITRK1*, and *IGSF11* play roles in synaptogenesis and learning flexibility.^33–35^ A notable feature of HG is heightened sensitivity to food, drink, sound, and smell that leads to a learned association between these stimuli and nausea and vomiting. The mechanism known as conditioned taste aversion, is associated with 5-HT content and dopamine signaling^36,37^ and *SLITRK1* knockout mice have serotonergic disturbances while *SYN3* regulates dopamine release.^35,38^ Common antiemetic medications used to treat HG target serotonin and dopamine receptors and vary in effectiveness from one patient to the next, so future study to determine whether genotype-phenotype associations related to medication effectiveness in HG patients may be worthwhile.^39^

Interestingly, 6 of the 10 candidate genes in this study *GDF15*, *GFRAL*, *IGFBP7*, *PGR*, *TCF7L2*, and *SYN3* have been associated with cachexia, a wasting condition with similar symptoms to HG including loss of appetite, weight loss, and muscle wasting.^10,40–45^ Manipulation of GDF15, GFRAL, IGFBP7, PGR, and TCF7L2 in animal models have exhibited effectiveness in reducing symptoms of cachexia.^41–45^ Thus, assuming analogous functions for these factors in HG, there is both genetic and biological support for causal and potentially reversible contributions for these genes in nausea and vomiting of pregnancy.

The association between HG and six genes (*GDF15*, *GFRAL*, *TCF7L2*, *IGSF11*, *FSHB*, *IGFBP7*) that are related to insulin and altered glucose metabolism,^46–51^ provides biological support for the theory that at least in some cases, glycemic status and insulin may be related to severity of NVP.^46^ It has been postulated that reduced energy intake in early pregnancy results in lower maternal insulin and IGF-1, tipping the scales toward an evolutionary advantage that favors augmentation of placental development and subsequent fetal growth. This theory is supported by animal husbandry practices such as those designed to stimulate placental development by placing ewes in poor pasture land in early pregnancy.^52^ Insulin also plays a role in conditioned taste aversion, so another theory is that perhaps these risk loci abnormally dysregulate learning and memory formation of aversive stimuli.^36^

Of note, a study of *TCF7L2* expression in mouse brain found neuronal expression in areas associated with sensing circulating nutrient levels including the area postrema.^53^ The diabetes-associated gene is a transcription factor that may control GLP-1 expression and is associated with liraglutide effects resulting in greater weight loss in obesity.^54^ A link between TCF7L2 and GLP-1 is intriguing because the GLP-1 receptor, GLP1R is expressed in the same cluster of neurons as GFRAL in the area postrema,^54,55^ and both GLP1R and GFRAL agonists are of great interest by pharmaceutical companies for their roles in appetite and weight loss.^6,56^ Understanding where TCF7L2 fits into this unfolding landscape is therefore of particular interest.

In addition to neuronal expression, *TCF7L2*, and 3 other candidate genes (*GDF15*, *IGFBP7*, and *PGR*), were found in this study to be differentially expressed during early placental development. *GDF15* and *TCF7L2* were primarily expressed in fetal derived trophoblasts during differentiation and invasion. If these genes play roles in placental development, there is evidence for redundancy as *TCF7L2* knockout mice lack a placental phenotype and are developmentally normal at birth, and murine and human *GDF15* knockouts are reportedly normal and fertile.^6,57–59^ Conversely, *IGFBP7* and *PGR* were primarily found in maternally derived spiral arteries, suggesting a maternal genetic contribution to spiral artery remodeling, and knockout of these genes in murine models contributes to a reduction in litter size and infertility, respectively.^60,61^

This study also identified individual associations between risk genes and adverse outcomes including preeclampsia, head circumference at birth, and birth weight. HG is associated with increased risk of preeclampsia^62^ and the *PGR* risk locus for HG (rs12790159) is in LD (r2=0.86, D’=-0.95) with the risk locus for PE (rs2508372).^63^ Associations between increased GDF15 levels at 30-34 weeks’ gestation and subsequent preterm preeclampsia have been noted previously,^64^ but to our knowledge, this is the first time a *GDF15* variant has been linked with serious preeclampsia. In addition, in utero exposure to HG is associated with smaller brains and neurodevelopmental delay at age 10,^58^ so the link to head circumference at birth and *IGSF11* and *PGR* loci for HG require further investigation.

A major purpose of genetic studies is to elucidate biological mechanisms associated with disease in the hopes that it will provide evidence to support the development of novel therapeutics that can benefit patients. The finding that genetic variation in *GDF15* is the most significant association in this study and in a recent GWAS of Japanese HG patients,^65^ and the identification of a genome-wide significant variant 2.8kb upstream of its receptor *GFRAL,* provide confidence in pursuing this therapeutic pathway. This study also predicted the *GDF15* risk locus to have opposing maternal and fetal contributions, consistent with our previous finding that lower circulating maternal GDF15 levels prior to pregnancy and higher circulating GDF15 levels produced by the fetal placenta during pregnancy are the main drivers of HG risk.^61^

Finally, in this study we found highly disproportionate conservation of risk loci which provides biological support for the theory that NVP evolved under strong selective pressure.^62^ Since approximately a third of pregnancies have no NVP symptoms, it may be an antiquated evolutionary mechanism no longer necessary for human survival. The 10 genetic associations provide intriguing avenues to advance our understanding and pursue novel therapeutic pathways for a common pregnancy condition that in its most severe form is associated with significant morbidity and even mortality for mothers and exposed offspring.

## METHODS

### Study cohorts and cohort-level GWAS

We included nine cohort level GWASs with varying sample sizes: 23andME (1,306 cases and 15,756 controls), Fejzo ExWAS (926 cases and 660 controls), FinnGen R8 (2,092 cases and 163,702 controls), Estonian Biobank (3,536 cases and 32,113 controls), UK Biobank (220 cases and 180,917 controls), Genes & Health (1,544 cases and 11,128 controls), HUNT (269 cases and 36,460 controls), eMERGE (504 cases and 6,364 controls), MoBa (577 cases and 14,361 controls). Together, these GWASs involved a total of 10,974 cases, 461,461 controls, and 43,961,852 SNPs. The population make-up of each study is shown in **Table 1**.

### Fejzo ExWAS

Exome-wide summary statistics from an EWAS on HG published previously were used in this study.^11^ Briefly, the source population for HG cases included patients treated with intravenous fluids for nausea and vomiting of pregnancy residing in the USA primarily recruited through advertising on the Hyperemesis Education and Research Foundation website (www.hyperemesis.org) from 2007 to 2017. Each case was asked to recruit a non-blood-related acquaintance with at least two pregnancies that went beyond 20 weeks of gestation to participate as a control. Whole-exome sequencing was performed by Regeneron Genetics Center (RGC, Tarrytown, NY, USA) with the Illumina NovaSeq platform using paired-end 75-bp reads (Illumina, San Diego, CA, USA). The captured bases were sequenced so that greater than 95% of samples passing initial quality control had at least 90% of the targeted bases covered at 20x or greater. Paired-end reads and genetic variants were called using the RGC DNAseq analysis pipeline. Ancestry was assigned using standard genetic prediction methods based on the intersection of SNPs between HapMap3 and the non-filtered project level VCF. SNPs were filtered to include common, high-quality SNPs and merged for the HapMap3 data set. All 926 cases and 660 control participants provided informed consent. The study was approved by the UCLA Institutional Review Board.

### Fejzo GWAS

We performed firth logistic regression with REGENIE^66^ on 451,677 variants while adjusting for 10 genotype PCs. We did not adjust for age because maternal age at time of affected/unaffected pregnancies could not be determined.

### UK Biobank GWAS

In the UK Biobank, cases included in the study were defined as >/= 1 live birth, excessive vomiting pregnancy. Controls were defined as >/= 1 live birth, no excessive vomiting pregnancy.

We performed firth logistic regression with the REGENIE package on 59,366,184 variants while adjusting for age, genotype platform, and 40 genotype PCs. 17,079,576 variants remained after removing variants with minor allele frequency <0.001 and INFO score ≤ 0.3.

### FinnGen GWAS

FinnGen is a public-private collaborative effort combining genotyping and digital health record data from Finnish health registries with. Herein we accessed publicly available summary statistics from release 8.^17^ Individuals in FinnGen were genotyped with Illumina and Affymetrix chip arrays with QC to remove samples and variants of poor quality. Genome-wide imputation was performed using reference Finnish whole-genome sequence data. HG cases (N=2,092) were defined using nationwide registries based on ICD codes. Females with no HG code and no other maternal disorders^67^ were included as controls (N=163,702). Sex, age, 10 PCs, and genotyping batch were included as covariates in the analysis. GWAS results for r8 were performed using REGENIE package. We filtered out association statistics for variants that exhibited an INFO score < 0.3 in any contributing substudy within FinnGen.

### 23andMe GWAS

Genome-wide summary statistics were provided from a previously published GWAS on HG.^65^ 23andMe, Inc. is a personal genetics company where customers can consent to participate as research subjects. All 1306 cases who reported via an online survey that they received IV therapy for NVP and 15,756 controls who reported no NVP provided informed consent and answered online surveys according to a human subjects’ protocol approved by Ethical & Independent Review Services, a private institutional review board. Cases and controls were genotyped on one of four custom Illumina genotyping arrays, and additional genotypes were imputed using the September 2013 release of the 1000 Genomes Project Phase 133 reference haplotypes. All participants were filtered to select for European ancestry. Logistic regression was applied using age and five genotype principal components as covariates.

### Estonian Biobank GWAS

The Estonian Biobank (EstBB) is a population-based biobank that includes over 200,000 participants, representing 20% of the total Estonian population. Details of EstBB genotyping procedure have been described in previous publications.^68,69^ In summary, all EstBB participants were genotyped using Illumina arrays at the Core Genotyping Lab of the Institute of Genomics, University of Tartu. Samples were imputed using a population specific imputation reference panel of 2,297 whole genome sequencing samples.^70^ For the association analysis, a total of 3536 cases and 32113 controls were included. Cases were defined as women having an ICD-10 code O21 and its subcodes, while controls were defined as women without O21 who had a history of delivery (identified by the presence of any delivery ICD-10 codes: O80, O81, O82, O83, O84). Briefly, REGENIE v2.0.4^66^ was used for analysis, with age and the first 10 principal component (PCs) used as covariates.

### Genes & Health GWAS

Genes & Health is a longitudinal population-based study of adult British-Bangladeshi and British-Pakistani individuals (SAS ethnicity) from East London, Bradford and Manchester. There are currently ∼60,000 participants. Data from the July 2021 data freeze was used, which includes 44,396 participants. See www.genesandhealth.org and the cohort profile.^16^ DNA was extracted from Oragene saliva samples and genotyped on the Illumina GSA3v chip and TOPMed-r2 imputed. These data are linked to extensive health record data from primary care, local secondary care, and national datasets. Cases were defined by ICD10 O21. Control pregnancies were defined as having one live birth without any NVP reported. REGENIE software was used to perform the GWAS using firth logistic regression (–firth approx).

### HUNT GWAS

The Trøndelag Health Study (HUNT) is a population-based health survey including four recruitment waves—HUNT1 (1984-1986), HUNT2 (1995-1997), HUNT3 (2006-2008) and HUNT4 (2017-2019)—concentrated in the North-Trøndelag area, where all adults > 20 years of age were invited to participate.^71^ Electronic health records from the Trøndelag county hospitals (Nord-Trøndelag Hospital Trust, including St. Olavs, Namsos, and Levanger Hospitals) hold International Classification of Diseases and Related Health Problems (ICD) codes back to 1987 and were last accessed August 8, 2021. DNA from 71,860 HUNT samples in HUNT2-3 was used in this study which were genotyped using one of three different Illumina HumanCoreExome arrays (HumanCoreExome12 v1.0, HumanCoreExome12 v1.1 and UM HUNT Biobank v1.0).^18^

ICD codes were grouped into phecodes.^72^ Phecode 643.1, “Hyperemesis gravidarum” includes one or more billing code 643.0, 643.00, 643.01, 643.03, 643.1, 643.10, 643.11 and 643.13 for ICD9 and O21.0, O21.1 for ICD10. SAIGE v0.43.1 was used with covariates birth year, genotyping batch, and the first four principal components from genotypes. Variants with MAC < 10 were excluded.

The genotyping in Trøndelag Health Study and work presented here was approved by the Regional Committee for Ethics in Medical Research, Central Norway (2018/1622). All participants signed informed consent for participation and the use of data in research.

### eMERGE GWAS

The Electronic Medical Record and Genomics (eMERGE) Network is a national network combining DNA biorepositories with electronic health record systems.^13^ It contains data from individuals at more than 10 sites across the U.S. Individuals from all eMERGE sites were genotyped using the Illumina 660WQuad array for those reported as European or unknown ancestry and the Illumina 1M-Duo array for those of African ancestry. Imputation and standard quality control procedures performed on each site cohort from eMERGE, including filtering based on sample quality and composition (e.g., sample genotyping call rate and relatedness) and marker quality (e.g., marker genotype call rate, concordance, and Hardy-Weinberg Equilibrium), as well as the additional quality control procedures used to merge genotype data from each site (e.g., strand orientation analysis).^73^ Individuals were included if they were females aged 18 years or older with International Classification of Disease, 9th and 10th revisions (ICD-9/ICD-10) codes indicating pregnancy and/or delivery. NVP/HG cases were defined as those with ICD-9 or ICD-10 codes for the conditions (643 and subcodes; O21 and subcodes); while controls were those without these codes. We performed firth logistic regression, adjusting 10 genotype PCs, using PLINK.^74^ Variants with info scores < 0.5 and minor allele frequencies less than 0.001 were excluded.

### MoBa GWAS

The Norwegian Mother, Father and Child Cohort Study (MoBa) is a population-based pregnancy cohort study conducted by the Norwegian Institute of Public Health.^75^ Participants were recruited from all over Norway from 1999-2008. The women consented to participation in 41% of the pregnancies. The cohort includes approximately 114,500 children, 95,200 mothers and 75,200 fathers. The Medical Birth Registry of Norway (MBRN) is a national health registry containing information about all births in Norway.

Genotyping data were acquired collectively by multiple Norwegian research centers on various genotyping platforms and are managed by the Norwegian Institute of Public Health (NIPH), information on the different genotyping batches are available from the NIPH (github.com/folkehelseinstituttet/mobagen), this study was conducted on the version 1.5 of the genotypes. Quality control of the genotypes was conducted by the PsychGen team, details on the pipeline are available in the respective publication^19^ and code repository (github.com/psychgen/MoBaPsychGen-QC-pipeline). The current study is based on version 10 of the participant questionnaire files. Pregnancies were classified as follows:

-No NVP: the mother did not report nausea or vomiting at any point of the pregnancy
-NVP: the mother reported nausea or vomiting but did not report being hospitalized for it
-HG: the mother reported being hospitalized due to prolonged nausea or vomiting

GWAS was conducted as a case-control using Regenie^66^ v. 3.2 as available from Bioconda^76^ and operated using SnakeMake^77^using Saddlepoint approximation against the genome of the mothers.

### GWAS meta-analysis

To enable coherent meta-analysis across all studies, we annotated datasets with Single Nucleotide Polymorphism Database release 155 (dbsnp155)^78^, filtered out SNPs with minor allele frequency ≥ 0.001 and INFO score ≤ 0.3, and synchronized datasets based on genome build GRCh38.^79^ We first lifted over 23andMe, Fejzo, FinnGen, Estonian Biobank, and UK Biobank from GRCh37 to GRCh38 using MungeSumstats.^80^ Then, we meta-analyzed 43,961,852 SNPs in 9 GWASes (23andMe, Fejzo, FinnGen, Estonian Biobank, UK Biobank, Genes & Health, HUNT, eMERGE, MoBa) using fixed effect analysis with METAL^81^ (version 2020-05-05) while allowing for heterogeneity. (**Supplementary Figure 2**).

### Fine mapping

We extracted SNPs that are present in both 515 unrelated Europeans individuals from the 1000 Genomes Project and meta-analysis summary statistics. SNPs with inconsistent allele coding between the two datasets were flipped based on 1000 Genomes Project. Ambiguous SNPs and SNPs with minor allele frequency below 0.01 were removed. 19,645 SNPs remained after.

We fine-mapped 10 regions within 1 Mb distance from the 10 lead SNPs in meta-analysis with minimum absolute correlation of 0.3 and coverage of 0.90 using susieR^20,82^ (version 0.12.35), a method that performs variable selection based on SuSiE (Sum of Single Effect) model and iterative Bayesian stepwise selection (IBSS) fitting procedure under a sparse Bayesian regression.

### Enrichment analysis

We used LDSC (version1.0.1) to perform partitioned LD score regression^83^ on meta-analysis summary statistics using 1000Genomes^84^ European Phase 3 baseline LD and HapMap3^85^ weights. Variants in the major histocompatibility complex (MHC) regions have been removed due to the uncommon LD structure. We estimated heritability and enrichments for 97 functional annotations while converting heritability from observed scale to liability scale (2.3% sample prevalence and 2% population prevalence).

### PheWAS

We conducted a phenome-wide association study on 10 variants identified by meta-analysis and around 3,380 phenotypes from 3 studies on Open Target Genetics^85,86^, including UK BioBank, FinnGen, and GWAS Catalog. The p-value threshold after the Bonferroni correction is 1.48×10^−6^.

### Look ups for selected pregnancy outcomes in MoBa

Phenotypes for maternal characteristics (maternal weight/weight gain/BMI at beginning of pregnancy, at 15 weeks’ gestation, and at term), pregnancy characteristics (preeclampsia, gestational diabetes, pregnancy duration, and placental weight), and offspring characteristics and outcomes (perinatal death, birth weight, head circumference) were obtained from the participant questionnaires and from the MBRN. Association of NVP/HG status with binary phenotypes was evaluated using Fisher’s exact test, association with linear phenotypes was evaluated using Student’s t-test. Association with the lead variants of the loci genome-wide significant in the meta-analysis was evaluated for each phenotype using logistic or linear regression for binary and numeric phenotypes, respectively. For each pregnancy, a risk score was computed using the effect size estimates of the lead variants in the meta-analysis and the genotypes of the mother. Association of the phenotypes with the risk scores was computed as for the variants. All analyses were conducted in R version 4.2.2.

### Spatiotemporal candidate gene expression during early placental development

Full details about cohort design and sample preparation have been published previously.^87^ Briefly, the study’s retrospective cohort design featured decidua tissue samples from elective pregnancy terminations at a single outpatient clinic. Thorough medical history was captured, and a board-certified gynecologist extracted specific patient details. The tissue management approach involved reviewing whole tissue sections to identify suitable samples containing decidual tissue and spiral arteries. Selected blocks were assembled into a Tissue Microarray (TMA) used in this study. Artery remodeling stage was manually determined by an expert annotator (E.S). The spatial transcriptomics experiment was performed using NanoString Technologies according to company manuals as published previously.^87^ Sample collection was performed as indicated in the GeoMx DSP instrument user manual (MAN-10088-03). Slides were loaded into the GeoMx DSP instrument and scanned. For each tissue sample, we selected regions of interest (ROI) corresponding to one of the following categories: artery (N=17), decidua (N=10), floating villi (N=12), VCT (N=15), interstitial EVT (N=23), anchoring EVT (N=20), endovascular EVT (N=8), endometrial glands (N=4) from 30 patients between 6-20 weeks’ gestation; Each ROI was collected into a single well in a 96-well plate. We performed library preparation per manufacturer instructions. Libraries were paired-end sequenced (2 × 75) on a NextSeq550 with up to 400 million total aligned reads. We gathered raw counts of each gene from each sample via the NanoString GeoMx NGS processing pipeline. We performed quality control aligned with NanoString’s data analysis manual, using the default parameters outlined there. We used recommended approach by the manufacturer for GeoMx data background subtraction using negative control probe reads and normalization. For normalization, we divided the counts of all genes. This entailed dividing the counts of all genes within a sample by that sample’s 75th percentile of expression, followed by a multiplication with identical scaling factors for all samples - determined by the geometric mean of all 75th percentiles. P-value calculation for differential expression between sample types was done using the Kruskal-Walis test.

### Conditioned parental and fetal effect estimates in MoBa

To estimate conditioned fetal, maternal, and paternal effect estimates, we conducted separate GWASes of NVP severity encoded as 0, 1, and 2 for no NVP, NVP, and HG,^10^ against the genomes of the children, mothers, and fathers in MoBa. NVP status and GWAS analyses were conducted as in the maternal GWAS used in the meta-analysis. Then, for the lead SNPs of the loci that were genome-wide significant in the meta-analysis, we applied a weighted linear model (WLM) to the summary statistics of the fetal, maternal, and paternal GWASes.^21^ Specifically, the model estimates partitioned fetal, maternal, and paternal effects from GWAS summary statistics equivalent to conditioned effects that would otherwise be obtained from a joint model given by,

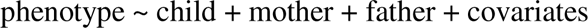

where phenotype here refers to NVP status; child, mother, and father the number of tested alleles in the genome of the child, mother, and father, respectively; and covariates the covariates used in the GWAS. The estimates of partitioned effects 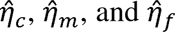 for the child, mother, and father, respectively, estimated from the GWAS estimates 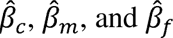 respectively, are:

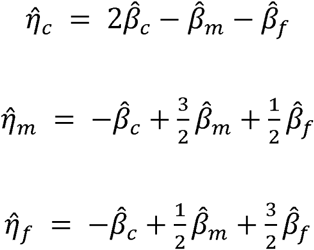

Standard errors for the effect estimates are then estimated as

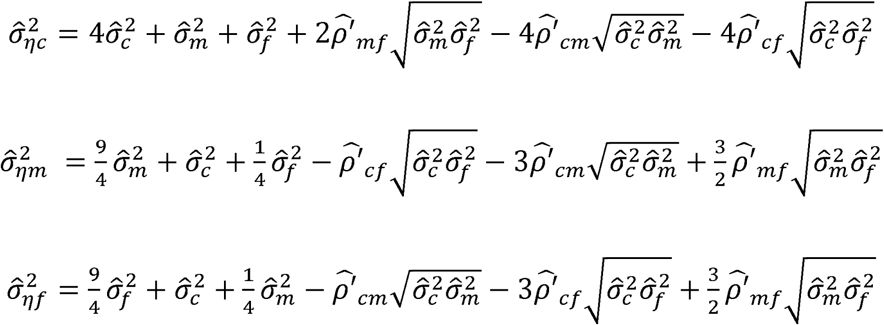

where 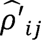 are coefficients accounting for sample overlap and correlation between 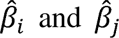 estimated from the intercept of a bivariate LD Score regression. P-values are calculated using a Z-test, with test statistic 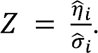^25,88^

Among the 10 association signals in the meta-analysis, 8 were found in the MoBa quality controlled genotypes, and one proxy association (rs112255166 r^2^=1 for rs76856932 nearest genes *TCF7L2-HABP2*) was identified among 2 missing SNPs using TopLD.^89^ Thus 9 SNPs were available for analysis.

### Maternal effects of individual SNPs by week of pregnancy

Mothers in MoBa reported whether they have suffered from NVP at different weeks of pregnancy (fhi.no/en/studies/moba/for-forskere-artikler/questionnaires-from-moba). To obtain cross sectional effect size estimates during pregnancy, we have conducted GWAS at these different time points using NVP as binary phenotype. GWASes were conducted as done in the maternal GWAS used in the meta-analysis.

### Analyses of GDF15 Processing

The 293-T cells were cultured in DMEM plus 10% fetal bovine serum and transfected with pCDNA3.1-GDF15, pCDNA3.1-GDF15 H202D or empty vector. Cells were washed three times with PBS and switched to serum-free DMEM 48 h after transfection. Conditioned media were harvested after 24 h. Proteins from conditioned medium were ethanol precipitated and subjected to SDS-PAGE. Western blotting was performed with αGDF15 rabbit polyclonal antibody recognizing GDF15 pro-peptide (Proteintech, Cat # 27455-1-AP). Processing efficiency was calculated by dividing the densitometry of pro-peptide by the densitometry of pro-peptide plus precursor in the conditioned medium samples. The experiments were repeated three times.

## Acknowledgements

We want to acknowledge the participants and investigators of 23andMe, Fejzo, FinnGen, Estonian Biobank, UK Biobank, Genes & Health, HUNT, eMERGE, and MoBa.

## Estonian Biobank

[Note: Estonian Biobank banner should refer to: Andres Metspalu, Lili Milani, Tõnu Esko, Reedik Mägi, Mari Nelis and Georgi Hudjashov Affiliation: Estonian Genome Centre, Institute of Genomics, University of Tartu (giving them credit for data collection, genotyping, QC and imputation) if a banner author is not allowed/available format in the journal, then add the banner in the Acknowledgements section. If an email address is necessary for the banner author, please use]

### Genes & Health

We thank Social Action for Health, Centre of The Cell, members of our Community Advisory Group, and staff who have recruited and collected data from volunteers. We thank the NIHR National Biosample Centre (UK Biocentre), the Social Genetic & Developmental Psychiatry Centre (King’s College London), Wellcome Sanger Institute, and Broad Institute for sample processing, genotyping, sequencing and variant annotation.

This work uses data provided by patients and collected by the NHS as part of their care and support. This research utilised Queen Mary University of London’s Apocrita HPC facility, supported by QMUL Research-IT.^90^

We thank: Barts Health NHS Trust, NHS Clinical Commissioning Groups (City and Hackney, Waltham Forest, Tower Hamlets, Newham, Redbridge, Havering, Barking and Dagenham), East London NHS Foundation Trust, Bradford Teaching Hospitals NHS Foundation Trust, Public Health England (especially David Wyllie), Discovery Data Service/Endeavour Health Charitable Trust (especially David Stables), Voror Health Technologies Ltd (especially Sophie Don), NHS England (for what was NHS Digital) - for GDPR-compliant data sharing backed by individual written informed consent.

Most of all we thank all of the volunteers participating in Genes & Health.

Genes & Health is/has recently been core-funded by Wellcome (WT102627, WT210561), the Medical Research Council (UK) (M009017, MR/X009777/1, MR/X009920/1), Higher Education Funding Council for England Catalyst, Barts Charity (845/1796), Health Data Research UK (for London substantive site), and research delivery support from the NHS National Institute for Health Research Clinical Research Network (North Thames). Genes & Health is/has recently been funded by Alnylam Pharmaceuticals, Genomics PLC; and a Life Sciences Industry Consortium of Astra Zeneca PLC, Bristol-Myers Squibb Company, GlaxoSmithKline Research and Development Limited, Maze Therapeutics Inc, Merck Sharp & Dohme LLC, Novo Nordisk A/S, Pfizer Inc, Takeda Development Centre Americas Inc.

We thank Social Action for Health, Centre of The Cell, members of our Community Advisory Group, and staff who have recruited and collected data from volunteers. We thank the NIHR National Biosample Centre (UK Biocentre), the Social Genetic & Developmental Psychiatry Centre (King’s College London), Wellcome Sanger Institute, and Broad Institute for sample processing, genotyping, sequencing and variant annotation.

This work uses data provided by patients and collected by the NHS as part of their care and support. This research utilised Queen Mary University of London’s Apocrita HPC facility, supported by QMUL Research-IT.^90^

We thank: Barts Health NHS Trust, NHS Clinical Commissioning Groups (City and Hackney, Waltham Forest, Tower Hamlets, Newham, Redbridge, Havering, Barking and Dagenham), East London NHS Foundation Trust, Bradford Teaching Hospitals NHS Foundation Trust, Public Health England (especially David Wyllie), Discovery Data Service/Endeavour Health Charitable Trust (especially David Stables), Voror Health Technologies Ltd (especially Sophie Don), NHS England (for what was NHS Digital) - for GDPR-compliant data sharing backed by individual written informed consent.

Most of all we thank all of the volunteers participating in Genes & Health.

### eMERGE Network (Phase III)

This phase of the eMERGE Network was initiated and funded by the NHGRI through the following grants: U01HG008657 (Group Health Cooperative/University of Washington); U01HG008685 (Brigham and Women’s Hospital); U01HG008672 (Vanderbilt University Medical Center); U01HG008666 (Cincinnati Children’s Hospital Medical Center); U01HG006379 (Mayo Clinic); U01HG008679 (Geisinger Clinic); U01HG008680 (Columbia University Health Sciences); U01HG008684 (Children’s Hospital of Philadelphia); U01HG008673 (Northwestern University); U01HG008701 (Vanderbilt University Medical Center serving as the Coordinating Center); U01HG008676 (Partners Healthcare/Broad Institute); U01HG008664 (Baylor College of Medicine); and U54MD007593 (Meharry Medical College).

### eMERGE Network (Phase IV)

This phase of the eMERGE Network was initiated and funded by the NHGRI through the following grants: U01HG011172 (Cincinnati Children’s Hospital Medical Center); U01HG011175 (Children’s Hospital of Philadelphia); U01HG008680 (Columbia University); U01HG011176 (Icahn School of Medicine at Mount Sinai); U01HG008685 (Mass General Brigham); U01HG006379 (Mayo Clinic); U01HG011169 (Northwestern University); U01HG011167 (University of Alabama at Birmingham); U01HG008657 (University of Washington Medical Center); U01HG011181 (Vanderbilt University Medical Center); U01HG011166 (Vanderbilt University Medical Center serving as the Coordinating Center). U01 HG 008657 (University of Washington)

### MoBa

We thank the Norwegian Institute of Public Health (NIPH) for maintaining and distributing MoBa data. The work was supported by the Research Council of Norway through its Centres of Excellence funding scheme, project number 262700. This research is part of the HARVEST collaboration, supported by the Research Council of Norway (#229624). We also thank the NORMENT Centre for providing genotype data, funded by the Research Council of Norway (#223273 and #229624), South East Norway Health Authorities and Stiftelsen Kristian Gerhard Jebsen. We further thank the Center for Diabetes Research, funded by the University of Bergen, the European Research Council (AdG #293574), Stiftelsen Kristian Gerhard Jebsen, Trond Mohn Foundation, the Research Council of Norway (#240413 and #301178), the Novo Nordisk Foundation (grant #54741), the Western Norway Health Authorities (grant #912250). The Norwegian Mother, Father and Child Cohort Study is supported by the Norwegian Ministry of Health and Care Services and the Ministry of Education and Research, NIH/NIEHS (contract no N01-ES-75558), NIH/NINDS (grant no.1 UO1 NS 047537-01 and grant no.2 UO1 NS 047537-06A1). Analyses on MoBa were performed using digital laboratories in HUNT Cloud at the Norwegian University of Science and Technology, Trondheim, Norway. We are grateful for outstanding support from the HUNT Cloud community. The participating families who contributed with data and biological material are gratefully acknowledged.

### Ethical agreements

The establishment of MoBa and initial data collection was based on a license from the Norwegian Data Protection Agency and approval from The Regional Committees for Medical and Health Research Ethics. The MoBa cohort is currently regulated by the Norwegian Health Registry Act. The current study was approved by The Regional Committees for Medical and Health Research Ethics (West committee number 2012/67 and South-East committee B number 2014/1178).

### HUNT

The Trøndelag Health Study (HUNT) is a collaboration between HUNT Research Center (Faculty of Medicine and Health Sciences, NTNU, Norwegian University of Science and Technology), Trøndelag County Council, Central Norway Regional Health Authority, and the Norwegian Institute of Public Health. The genotyping in HUNT was financed by the National Institutes of Health; University of Michigan; the Research Council of Norway; the Liaison Committee for Education, Research and Innovation in Central Norway; and the Joint Research Committee between St Olavs hospital and the Faculty of Medicine and Health Sciences, NTNU. The HUNT data may be accessed by application to the HUNT Research Centre.

## Data availability

Data from the Norwegian Mother, Father and Child Cohort Study and the Medical Birth Registry of Norway used in this study are managed by the national health register holders in Norway (Norwegian Institute of public health) and can be made available to researchers, provided approval from the Regional Committees for Medical and Health Research Ethics (REC), compliance with the EU General Data Protection Regulation (GDPR) and approval from the data owners. The consent given by the participants does not open for storage of data on an individual level in repositories or journals. Researchers who want access to data sets for replication should apply through helsedata.no. Access to data sets requires approval from The Regional Committee for Medical and Health Research Ethics in Norway and an agreement with MoBa.

The code used to analyze MoBa data along with its documentation is available at github.com/mvaudel/hyperemesis_gravidarum

## Contribution USC TEAM

M.S.F. conceived and supervised execution of the study as a whole, analyzed and interpreted results, and drafted the manuscript. N.M. and C.S. supervised the meta-analysis and contributed to study design and manuscript preparation. X.W. performed the meta-analysis and contributed to manuscript preparation.

## MoBa team

N.P.G. performed association analyses for EstBB data, N.P.G. and T.L. contributed to writing and reviewing the paper. Estonian Biobank Research Team collected and provided EstBB data.

I.A. analysed and interpreted NanoString data, E.S. performed NanoString experiments, M.A. supervised the study. YG performed experiments and data analysis. GG reviewed the manuscript.

JZ and DAvH were involved in the generation of the GWAS data for G&H. SF helped supervise the G&H work, and SF and DAvH supervised the collection of the G&H data.

E.A.J. performed analyses and reviewed and revised the manuscript.

L.B. lead analysist HUNT, K.H. and B.M.B. co-PI HUNT. B.M.B. lead contact HUNT.

Y.L. data contribution (e.g., phenotyping for eMERGE cohort that is used by this study) and manuscript review.

---

**Supplementary Figure S1.**
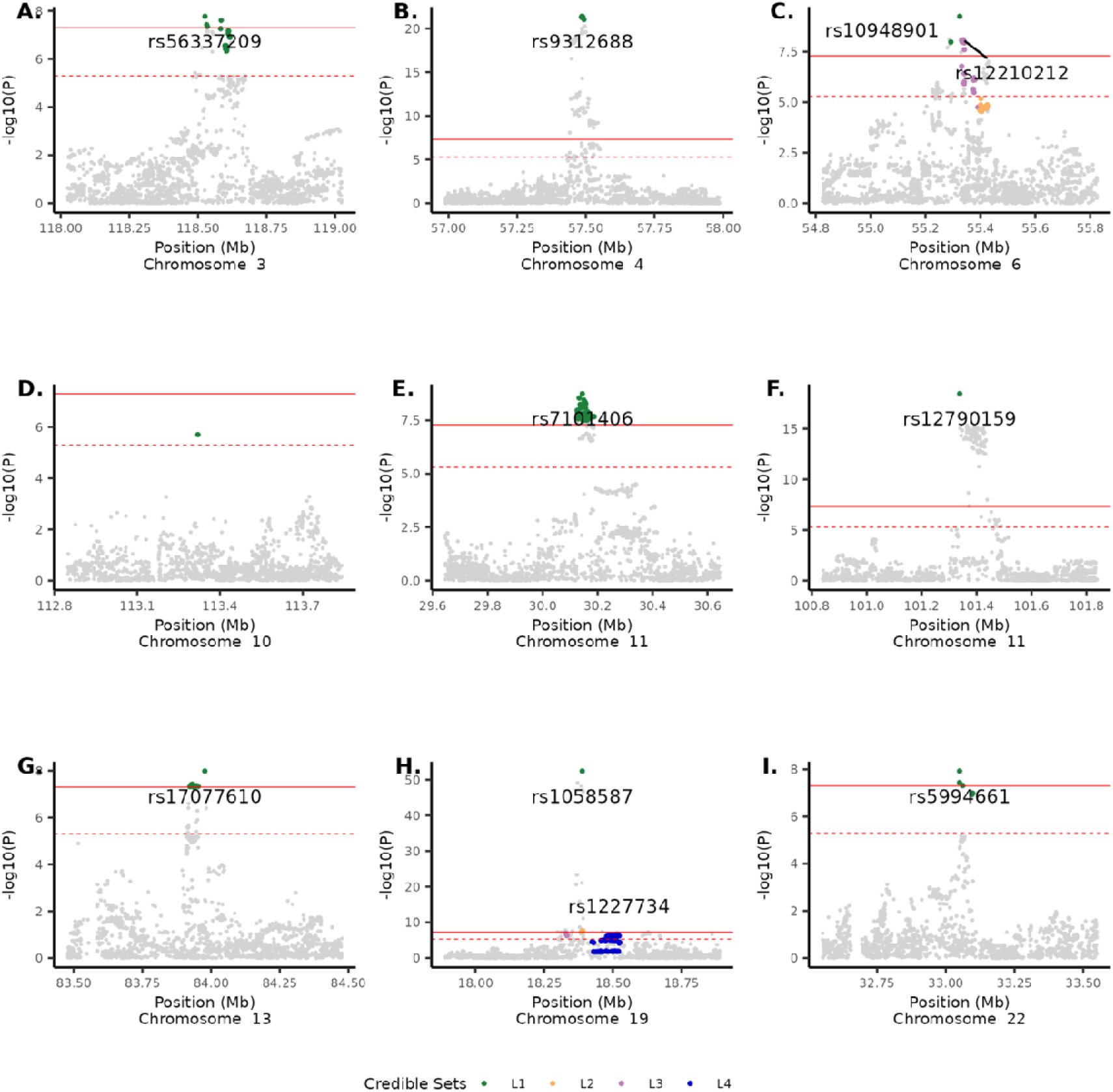
Locus zoom plots of 10 regions within 1 Mb distance from the 9 lead SNPs in meta-analysis with minimum absolute correlation of 0.3 and coverage of 0.90. One region failed to pass a relaxed genome-wide significant threshold (P <) was removed. **A.** chr3:118.0-119.0 Mb **B.** chr4: 57.0-58.0 Mb **C.** chr6:54.8-55.8 Mb **D.** chr10:112.8-113.8 Mb **E.** chr11:29.6-34.6 Mb **F.** chr11:100.8-110.8 Mb **G.** chr13:83.5-84.5 Mb **H.** chr19:17.9-18.9 Mb **I.** chr22:32.5-33.5 Mb.

**Supplementary Figure S2.**
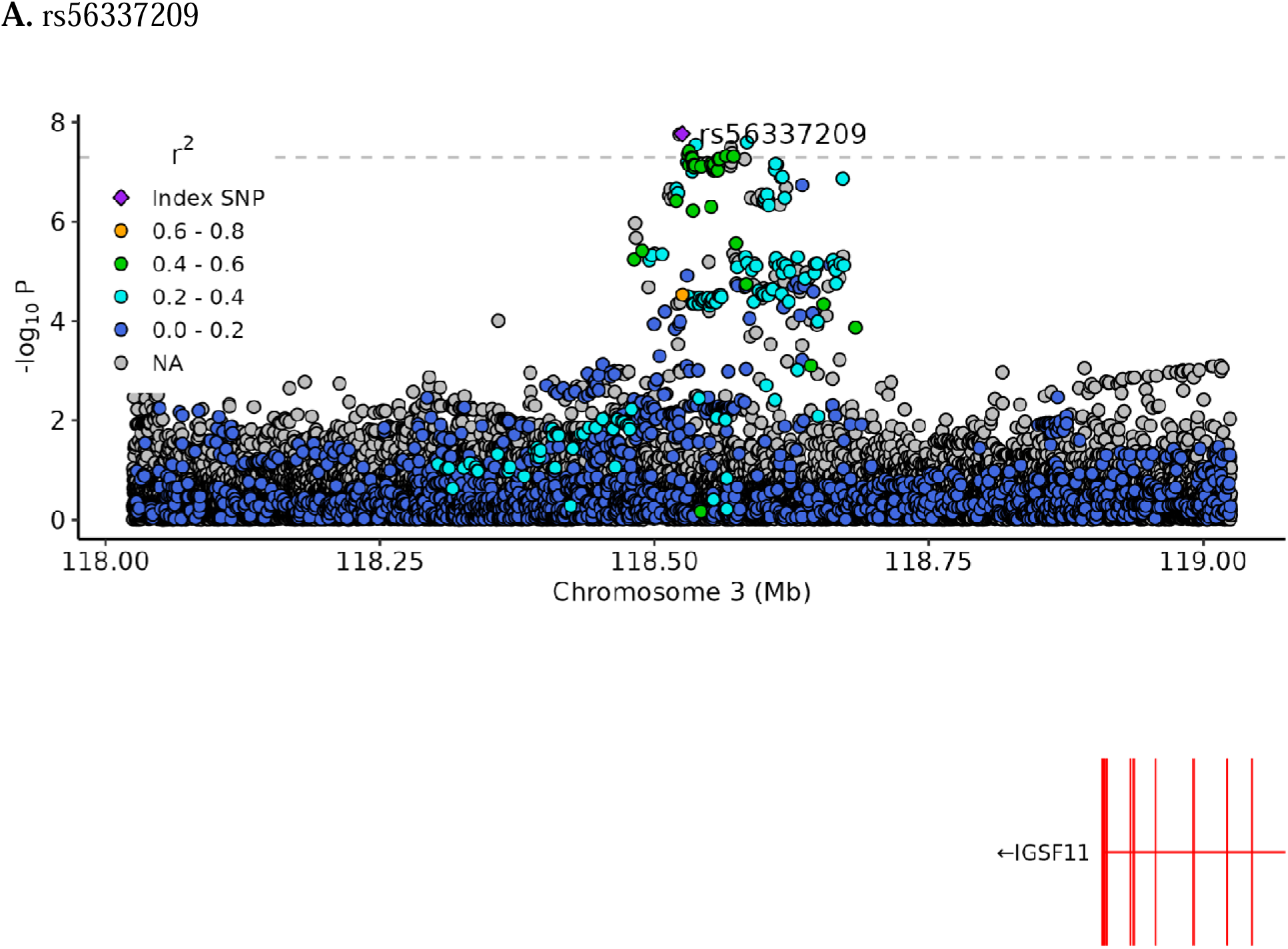
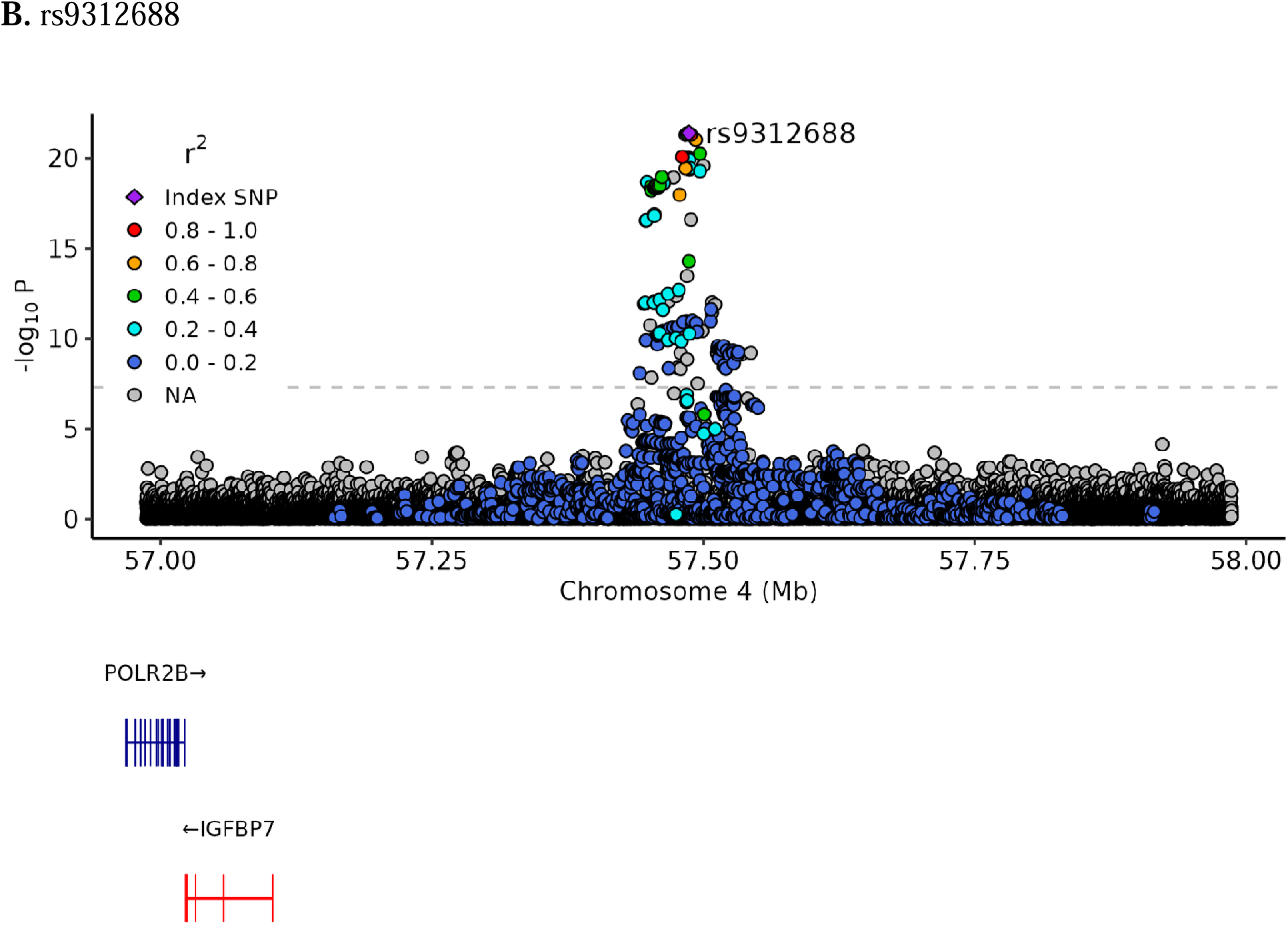
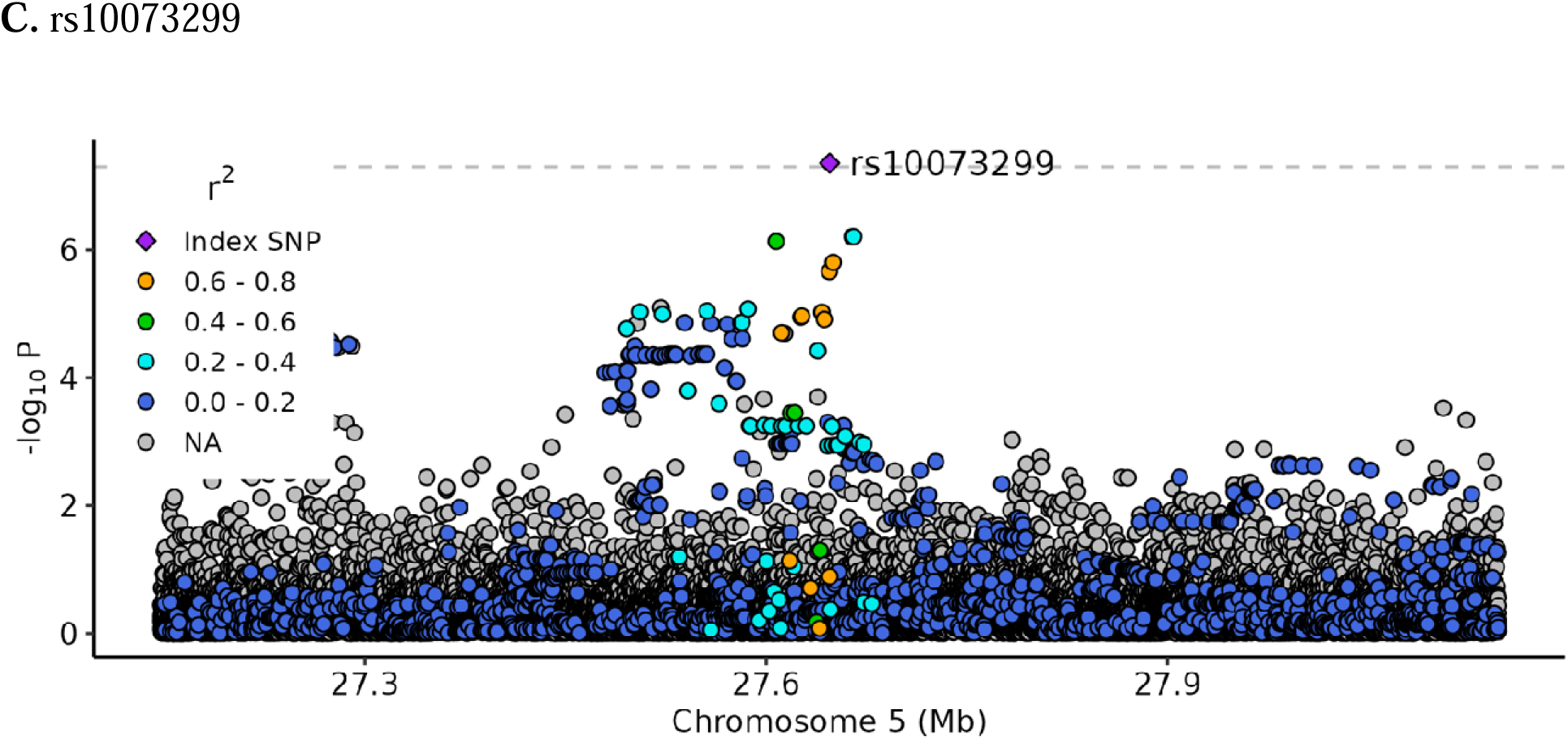
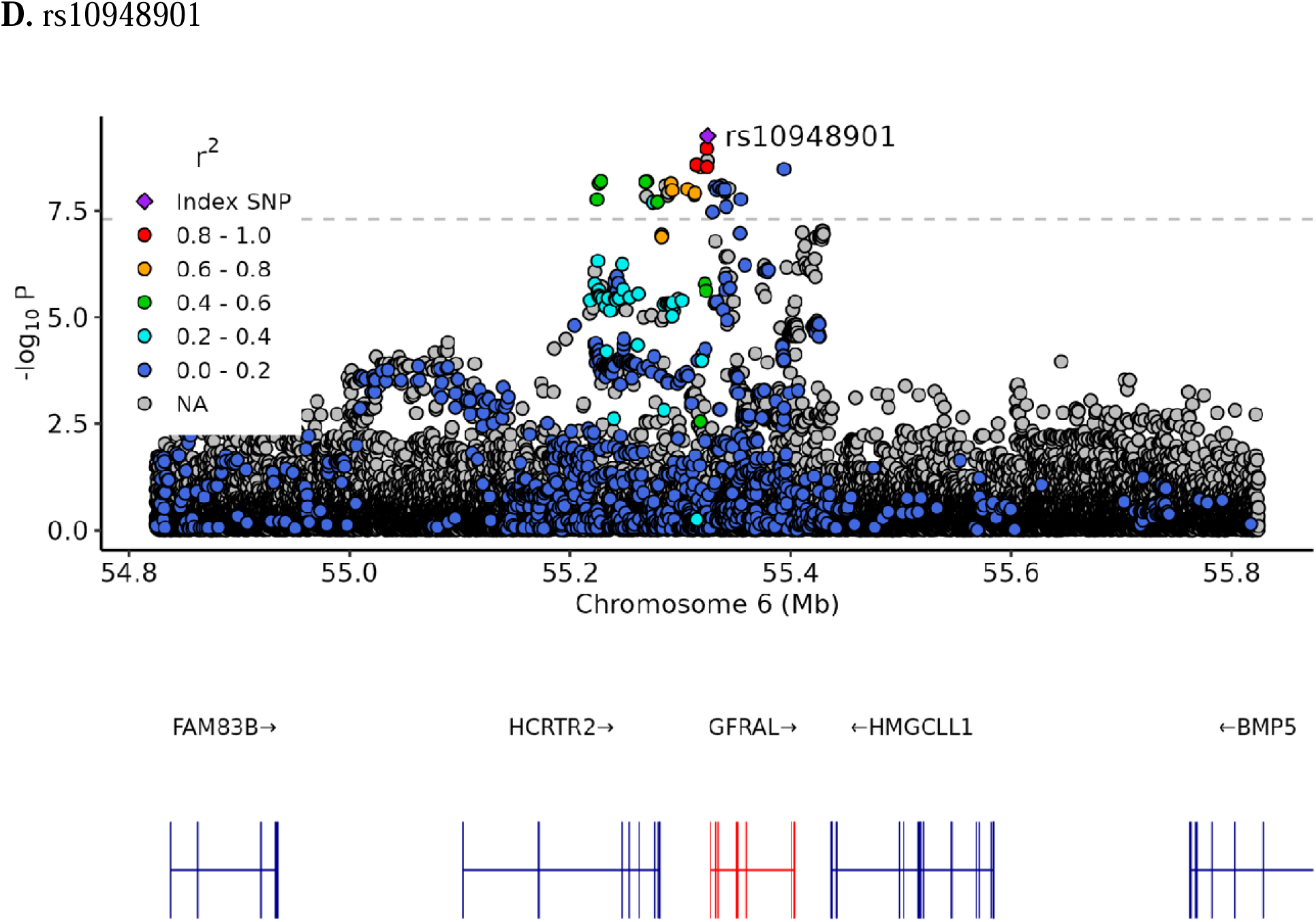
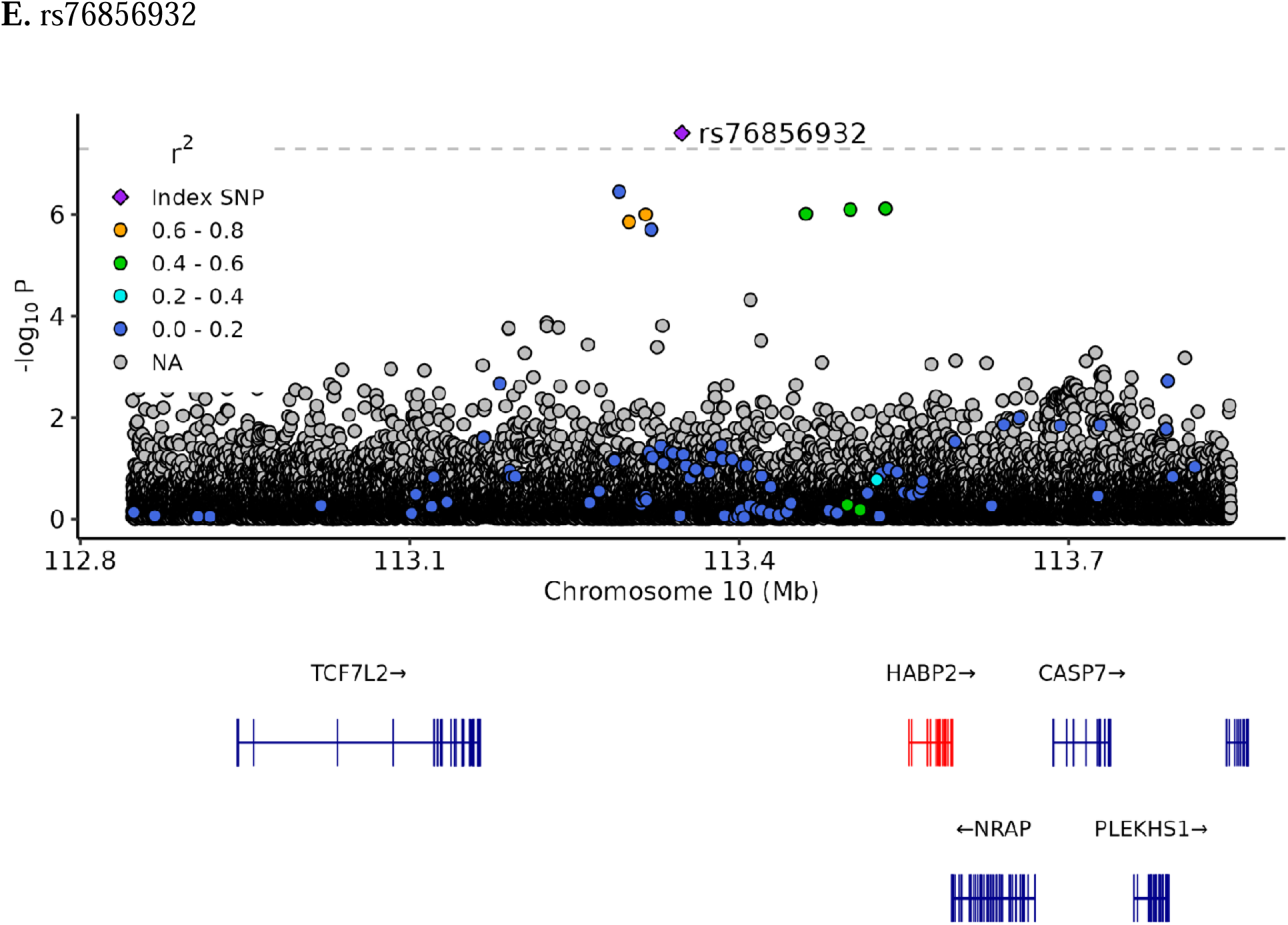
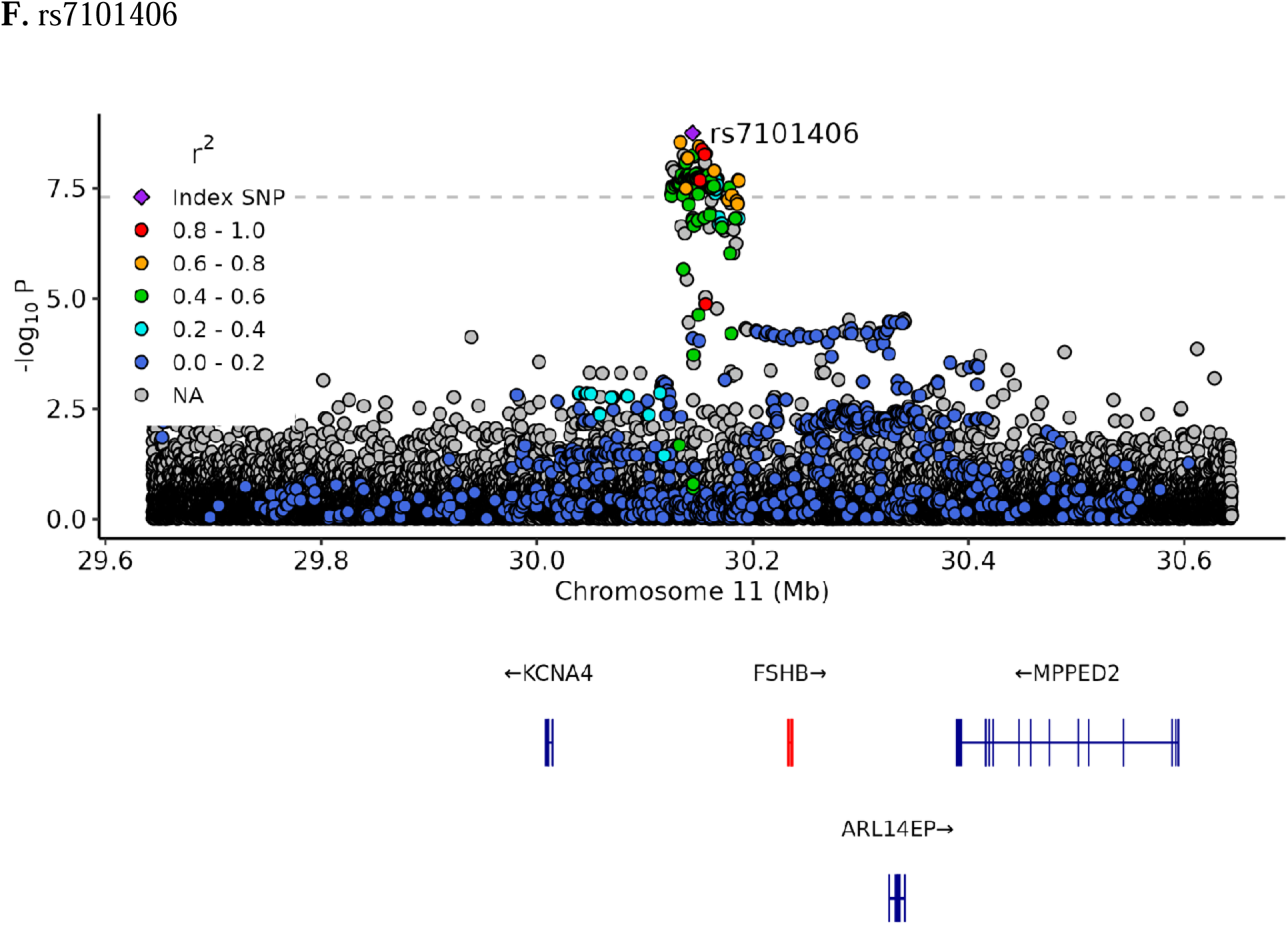
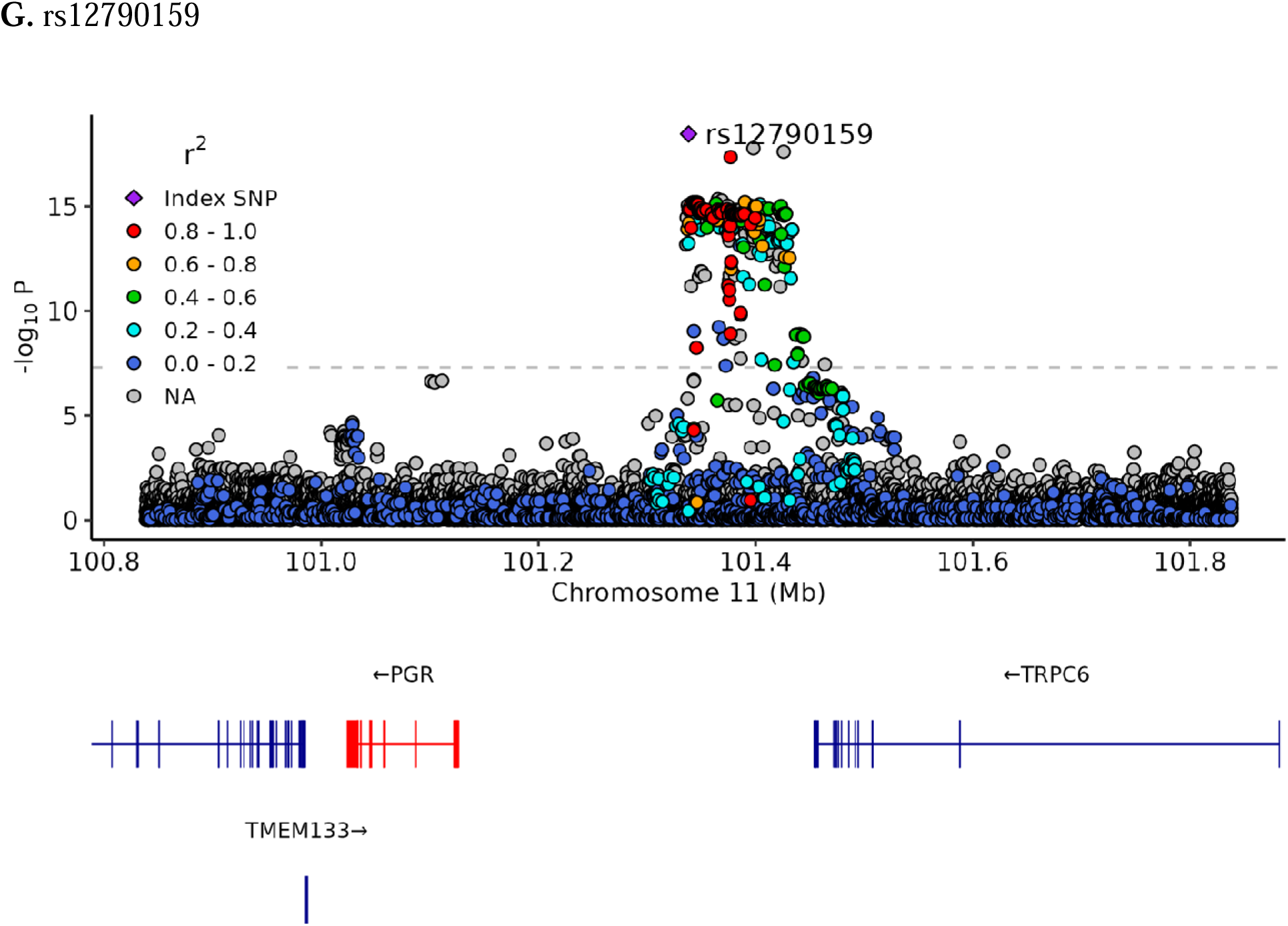
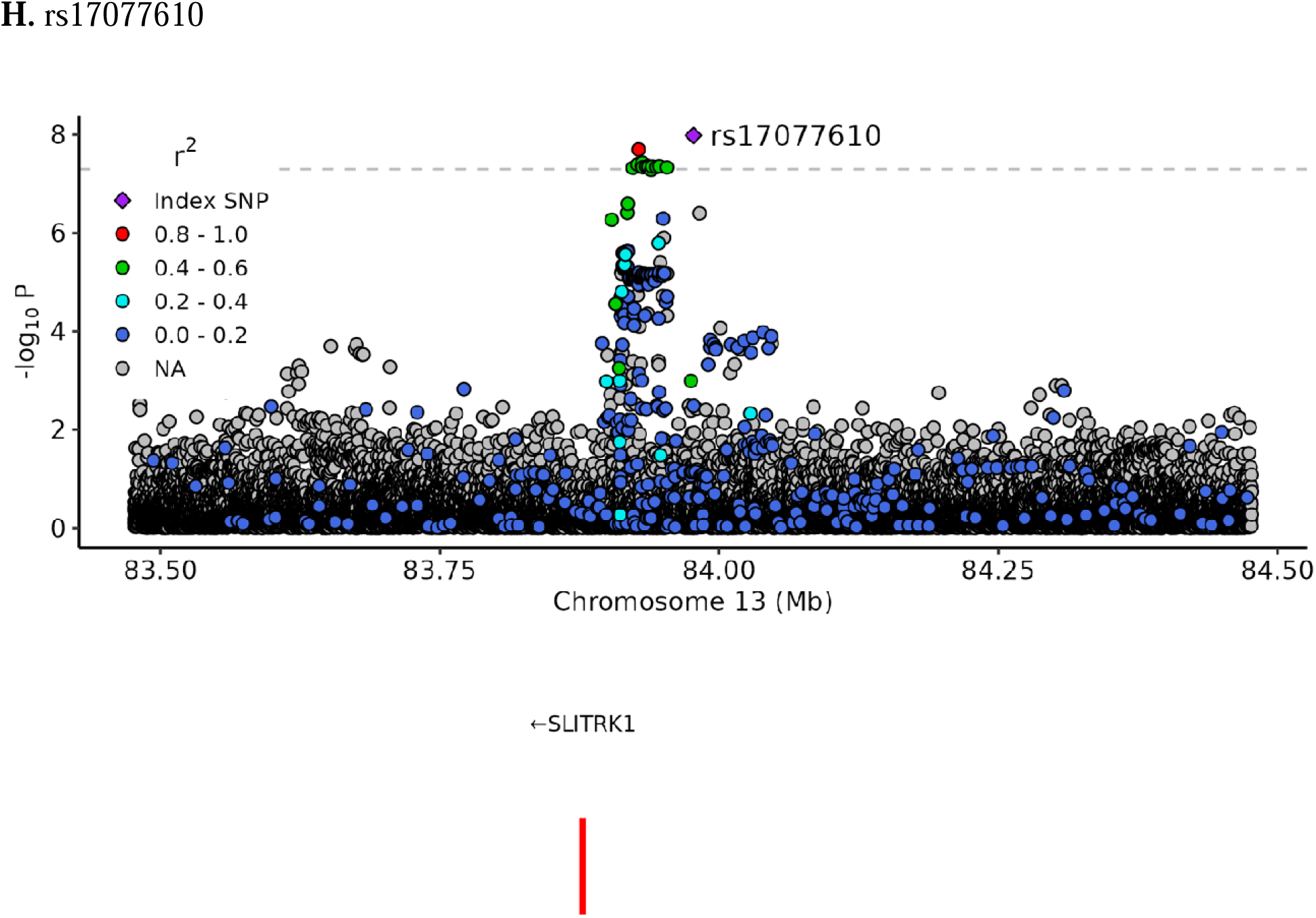
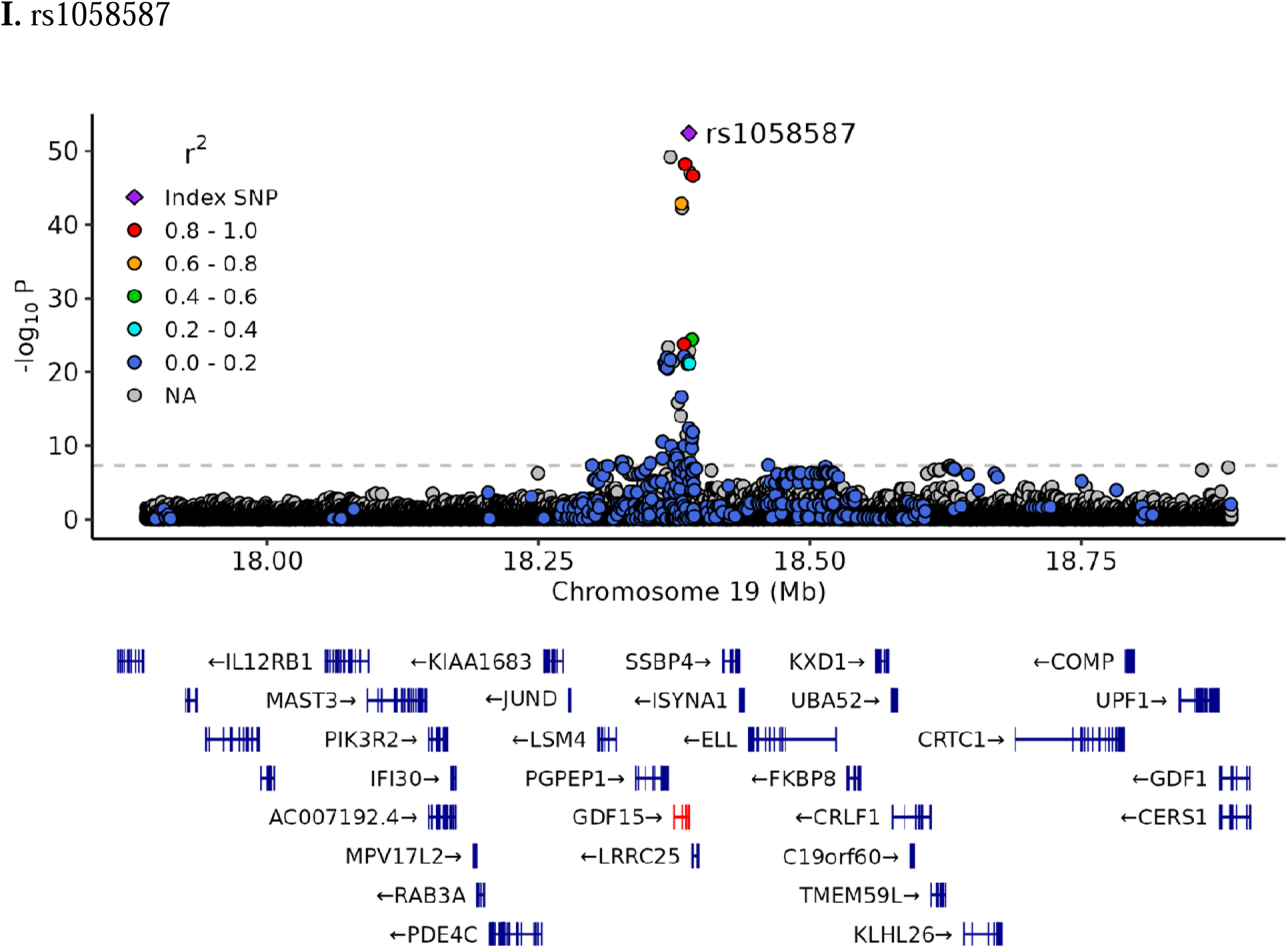
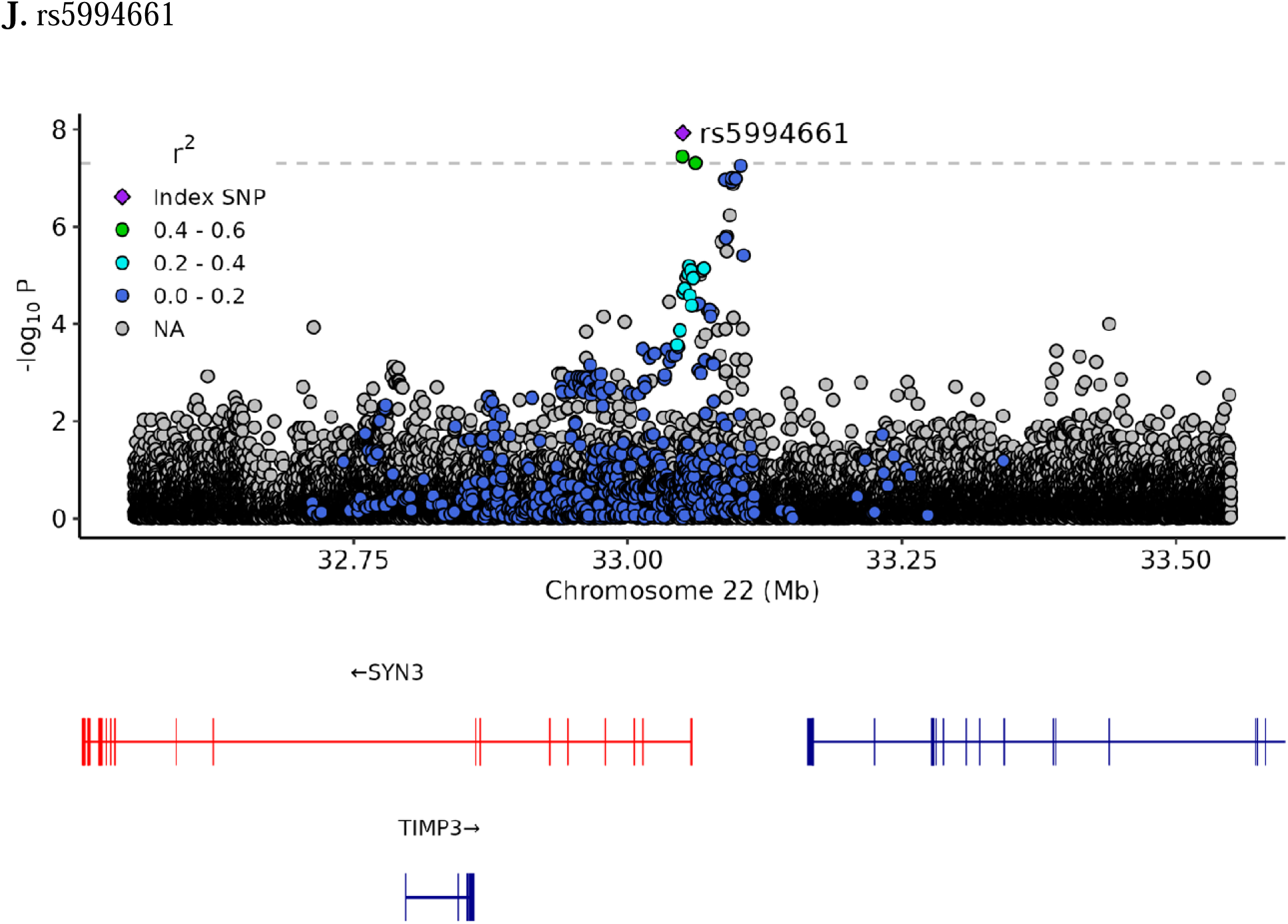
Regional association plots for 10 SNPs found in the meta-analysis. **A.** rs56337209 **B.** rs9312688 **C.** rs10073299 **D.** rs10948901 **E.** rs76856932 **F.** rs7101406 **G.** rs12790159 **H.** rs17077610 **I.** rs1058587 **J.** rs5994661.

**Supplementary Figure S3.**
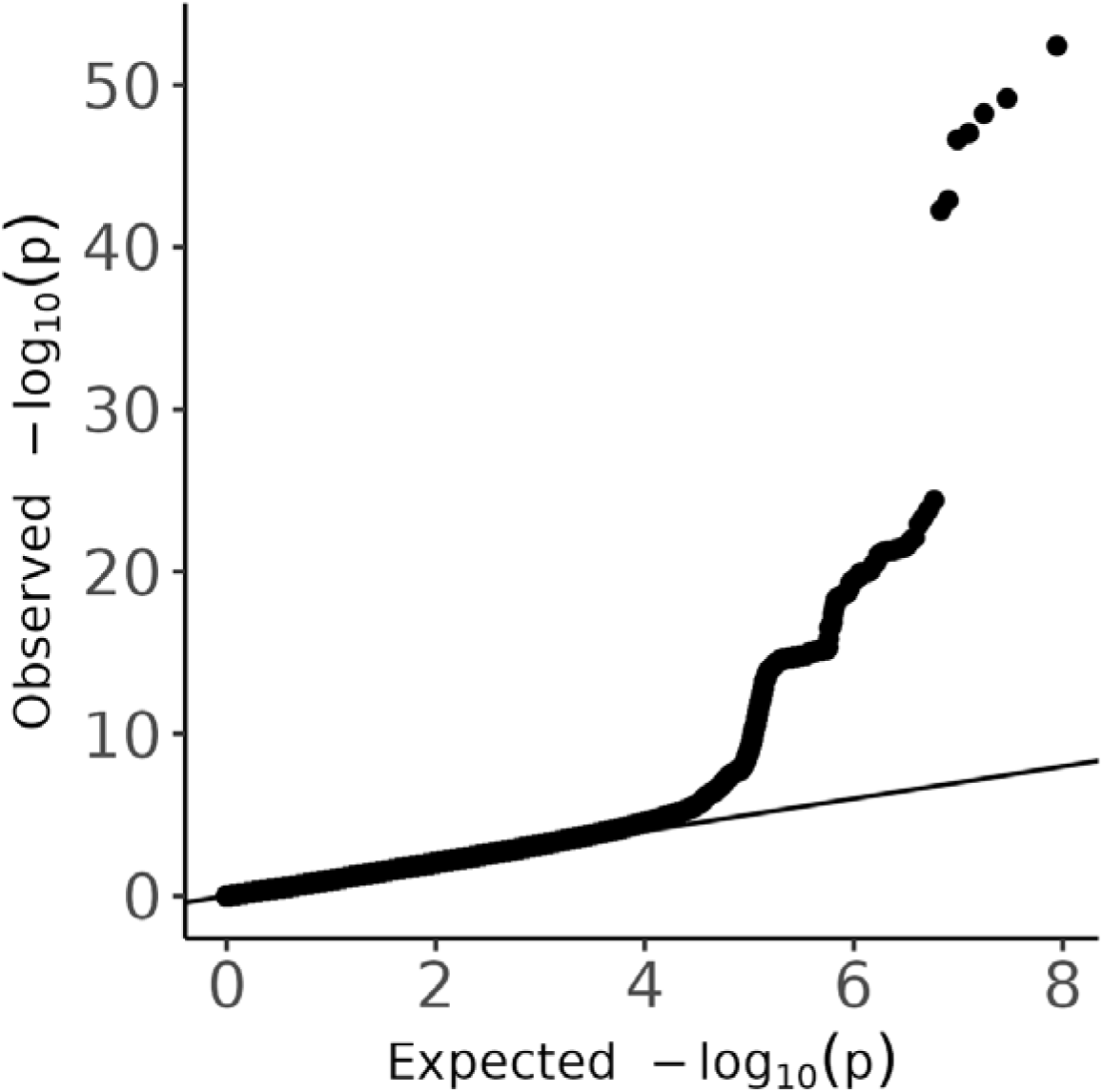
QQ Plot.

**Supplementary Figure S4.**
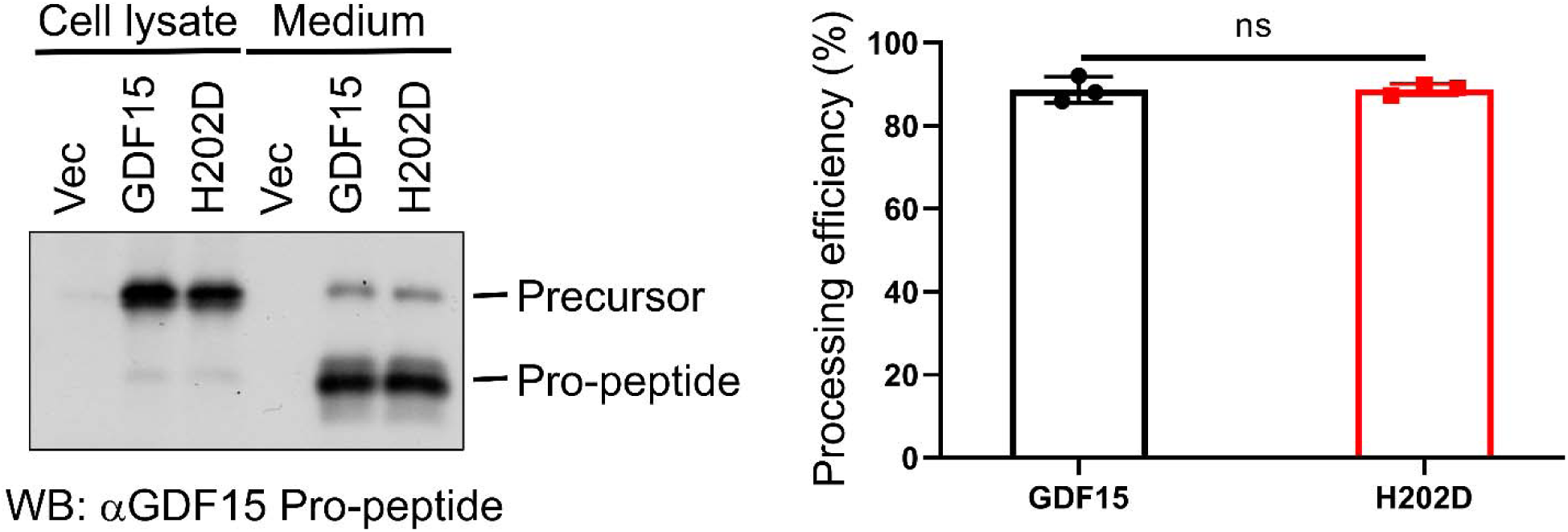
No significant difference in pro-GDF15 maturation between H (GDF15) and D (H202D) for rs1058587.

