## SUPPLEMENTAL TABLES 1-6 for "Multi-ancestry GWAS of severe pregnancy nausea and vomiting identifies risk loci associated with appetite, insulin signaling, and brain plasticity"

SUPPLEMENTARY TABLES S1-6

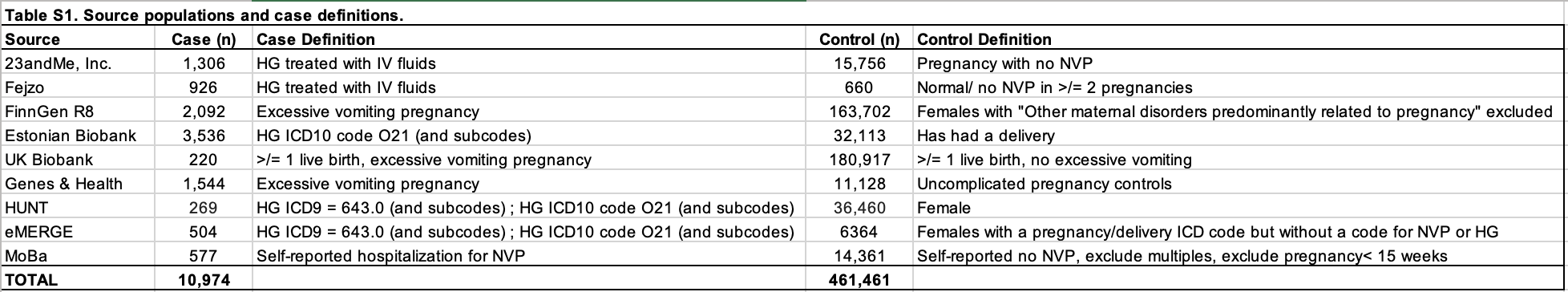

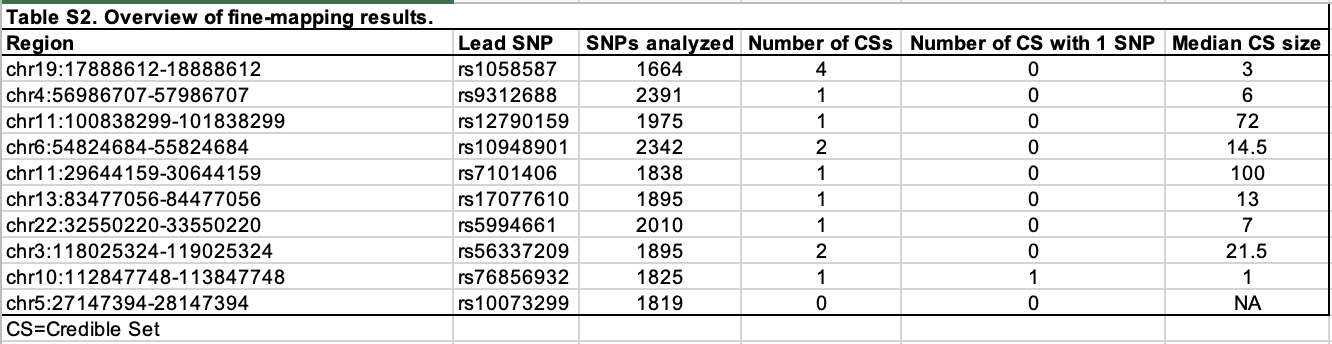

| **Table S3. Posterior Inclusion Probabilities (PIPs) for SNPs with non-zero effects in each credible set.** | | | | | |
| --- | --- | --- | --- | --- | --- |
| **SNP** | **CHR** | **POS** | **P** | **PIP** | L |
| rs523207 | 10 | 113320016 | 1.98E-06 | 0.92786978 | L1 |
| rs10895102 | 11 | 101365237 | 4.60E-16 | 0.04256553 | L1 |
| rs2508362 | 11 | 101390067 | 6.21E-16 | 0.03271108 | L1 |
| rs7122352 | 11 | 101363528 | 7.27E-16 | 0.02698624 | L1 |
| rs7121801 | 11 | 101403945 | 7.89E-16 | 0.0248407 | L1 |
| rs11224748 | 11 | 101394775 | 8.96E-16 | 0.02398927 | L1 |
| rs1943772 | 11 | 101374117 | 9.34E-16 | 0.02503667 | L1 |
| rs2155047 | 11 | 101339866 | 9.40E-16 | 0.02473805 | L1 |
| rs3018696 | 11 | 101395168 | 9.62E-16 | 0.02027661 | L1 |
| rs11224754 | 11 | 101423373 | 9.64E-16 | 0.02150243 | L1 |
| rs2508365 | 11 | 101400259 | 9.89E-16 | 0.02230263 | L1 |
| rs2513170 | 11 | 101400509 | 9.98E-16 | 0.02230263 | L1 |
| rs2508372 | 11 | 101397592 | 1.02E-15 | 0.01961416 | L1 |
| rs2513172 | 11 | 101399791 | 1.05E-15 | 0.02156595 | L1 |
| rs2508369 | 11 | 101399133 | 1.05E-15 | 0.02095977 | L1 |
| rs744021 | 11 | 101407294 | 1.20E-15 | 0.02171402 | L1 |
| rs1938991 | 11 | 101412162 | 1.31E-15 | 0.01967198 | L1 |
| rs7926811 | 11 | 101420752 | 1.31E-15 | 0.02032526 | L1 |
| rs9735005 | 11 | 101395454 | 1.37E-15 | 0.01899107 | L1 |
| rs12806321 | 11 | 101373798 | 1.38E-15 | 0.01909002 | L1 |
| rs1943771 | 11 | 101374100 | 1.41E-15 | 0.01909002 | L1 |
| rs2513149 | 11 | 101386424 | 1.41E-15 | 0.01827864 | L1 |
| rs10895101 | 11 | 101365164 | 1.46E-15 | 0.0179657 | L1 |
| rs1938978 | 11 | 101347728 | 1.46E-15 | 0.01833484 | L1 |
| rs10791464 | 11 | 101395349 | 1.47E-15 | 0.0178061 | L1 |
| rs4430478 | 11 | 101382155 | 1.50E-15 | 0.01527306 | L1 |
| rs12789640 | 11 | 101367751 | 1.52E-15 | 0.01739639 | L1 |
| rs10750601 | 11 | 101377222 | 1.59E-15 | 0.01438714 | L1 |
| rs10736618 | 11 | 101377122 | 1.60E-15 | 0.01396362 | L1 |
| rs12803803 | 11 | 101422837 | 1.64E-15 | 0.01442581 | L1 |
| rs35768873 | 11 | 101373623 | 1.72E-15 | 0.01631481 | L1 |
| rs11224752 | 11 | 101421879 | 1.72E-15 | 0.01389073 | L1 |
| rs36009736 | 11 | 101373615 | 1.73E-15 | 0.01582221 | L1 |
| rs11224753 | 11 | 101423242 | 1.73E-15 | 0.01360672 | L1 |
| rs10791465 | 11 | 101395357 | 1.75E-15 | 0.01477788 | L1 |
| rs7926045 | 11 | 101368393 | 1.75E-15 | 0.01533907 | L1 |
| rs1938972 | 11 | 101369400 | 1.76E-15 | 0.01533907 | L1 |
| rs1938975 | 11 | 101369145 | 1.77E-15 | 0.01565923 | L1 |
| rs7929078 | 11 | 101368559 | 1.80E-15 | 0.01533907 | L1 |
| rs2513165 | 11 | 101377939 | 1.83E-15 | 0.01279165 | L1 |
| rs10895103 | 11 | 101377687 | 1.89E-15 | 0.01243167 | L1 |
| rs1938976 | 11 | 101368889 | 2.00E-15 | 0.01400288 | L1 |
| rs2508375 | 11 | 101394148 | 2.03E-15 | 0.01386348 | L1 |
| rs7945321 | 11 | 101372802 | 2.09E-15 | 0.01359269 | L1 |
| rs10750602 | 11 | 101377317 | 2.16E-15 | 0.01175408 | L1 |
| rs2508360 | 11 | 101386673 | 2.17E-15 | 0.01380803 | L1 |
| rs12802904 | 11 | 101427705 | 2.18E-15 | 0.01322108 | L1 |
| rs7948518 | 11 | 101372913 | 2.18E-15 | 0.01319893 | L1 |
| rs2513166 | 11 | 101377832 | 2.19E-15 | 0.01112923 | L1 |
| rs2513178 | 11 | 101394194 | 2.23E-15 | 0.01347756 | L1 |
| rs1938981 | 11 | 101380917 | 2.24E-15 | 0.01083528 | L1 |
| rs35452558 | 11 | 101427029 | 2.24E-15 | 0.01285054 | L1 |
| rs12222432 | 11 | 101378681 | 2.25E-15 | 0.01083528 | L1 |
| rs2513158 | 11 | 101380402 | 2.26E-15 | 0.01083528 | L1 |
| rs1938987 | 11 | 101389559 | 2.29E-15 | 0.01301914 | L1 |
| rs2513164 | 11 | 101378142 | 2.30E-15 | 0.01112923 | L1 |
| rs12803811 | 11 | 101427816 | 2.31E-15 | 0.0124945 | L1 |
| rs12796333 | 11 | 101425227 | 2.32E-15 | 0.01239538 | L1 |
| rs12798288 | 11 | 101427325 | 2.35E-15 | 0.0124945 | L1 |
| rs2513154 | 11 | 101382789 | 2.35E-15 | 0.01301914 | L1 |
| rs12795968 | 11 | 101427317 | 2.37E-15 | 0.01182363 | L1 |
| rs2508353 | 11 | 101377786 | 2.37E-15 | 0.01083528 | L1 |
| rs1943773 | 11 | 101374529 | 2.44E-15 | 0.01080832 | L1 |
| rs2508352 | 11 | 101379653 | 2.53E-15 | 0.01002147 | L1 |
| rs2513146 | 11 | 101387512 | 2.55E-15 | 0.01196212 | L1 |
| rs11224756 | 11 | 101424188 | 2.56E-15 | 0.01182363 | L1 |
| rs2513155 | 11 | 101381900 | 2.58E-15 | 0.01080832 | L1 |
| rs10791461 | 11 | 101382510 | 2.59E-15 | 0.01195024 | L1 |
| rs2508358 | 11 | 101385058 | 2.60E-15 | 0.01195024 | L1 |
| rs2513151 | 11 | 101385423 | 2.60E-15 | 0.01195024 | L1 |
| rs2513162 | 11 | 101378628 | 2.61E-15 | 0.01002147 | L1 |
| rs2508357 | 11 | 101384261 | 2.67E-15 | 0.01195024 | L1 |
| rs2513167 | 11 | 101377824 | 2.69E-15 | 0.01080832 | L1 |
| rs7101406 | 11 | 30144159 | 1.80E-09 | 0.06495137 | L1 |
| rs35078732 | 11 | 30132930 | 2.87E-09 | 0.0370531 | L1 |
| rs3847634 | 11 | 30150047 | 3.64E-09 | 0.02952195 | L1 |
| rs10501125 | 11 | 30153120 | 4.25E-09 | 0.02540328 | L1 |
| rs1340032 | 11 | 30155332 | 4.96E-09 | 0.02451758 | L1 |
| rs3858481 | 11 | 30156270 | 5.23E-09 | 0.02401021 | L1 |
| rs3858479 | 11 | 30155463 | 5.44E-09 | 0.02204649 | L1 |
| rs637542 | 11 | 30144844 | 5.65E-09 | 0.0215331 | L1 |
| rs2082917 | 11 | 30136370 | 5.66E-09 | 0.02037315 | L1 |
| rs2226016 | 11 | 30154159 | 5.72E-09 | 0.02128105 | L1 |
| rs2460713 | 11 | 30146040 | 6.14E-09 | 0.01983121 | L1 |
| rs4285851 | 11 | 30139617 | 6.63E-09 | 0.01626076 | L1 |
| rs3847631 | 11 | 30139001 | 6.70E-09 | 0.01626076 | L1 |
| rs12796113 | 11 | 30137758 | 8.20E-09 | 0.01518738 | L1 |
| rs12794770 | 11 | 30137832 | 8.34E-09 | 0.01518738 | L1 |
| rs7113720 | 11 | 30155720 | 8.40E-09 | 0.01556582 | L1 |
| rs4923621 | 11 | 30125738 | 1.08E-08 | 0.01089483 | L1 |
| rs72885907 | 11 | 30164303 | 1.28E-08 | 0.01032722 | L1 |
| rs979210 | 11 | 30127324 | 1.31E-08 | 0.01014933 | L1 |
| rs10835625 | 11 | 30148214 | 1.45E-08 | 0.00890577 | L1 |
| rs11030973 | 11 | 30147198 | 1.50E-08 | 0.00860503 | L1 |
| rs3886880 | 11 | 30139254 | 1.56E-08 | 0.00739215 | L1 |
| rs1561088 | 11 | 30134472 | 1.57E-08 | 0.0088121 | L1 |
| rs78026352 | 11 | 30141228 | 1.60E-08 | 0.00713774 | L1 |
| rs3847637 | 11 | 30158211 | 1.63E-08 | 0.00793995 | L1 |
| rs3858484 | 11 | 30158402 | 1.66E-08 | 0.00767114 | L1 |
| rs12786317 | 11 | 30157656 | 1.67E-08 | 0.00767114 | L1 |
| rs3858480 | 11 | 30155573 | 1.69E-08 | 0.00765593 | L1 |
| rs72884326 | 11 | 30141197 | 1.72E-08 | 0.00665568 | L1 |
| rs10835626 | 11 | 30152416 | 1.73E-08 | 0.0069197 | L1 |
| rs7948109 | 11 | 30151770 | 1.76E-08 | 0.00668646 | L1 |
| rs3858482 | 11 | 30156274 | 1.78E-08 | 0.0069197 | L1 |
| rs10835629 | 11 | 30152891 | 1.80E-08 | 0.00668646 | L1 |
| rs2117911 | 11 | 30133943 | 1.80E-08 | 0.00642739 | L1 |
| rs12802602 | 11 | 30139361 | 1.80E-08 | 0.00642739 | L1 |
| rs10835618 | 11 | 30134282 | 1.81E-08 | 0.00620717 | L1 |
| rs11030972 | 11 | 30147017 | 1.83E-08 | 0.00646132 | L1 |
| rs12282309 | 11 | 30151640 | 1.84E-08 | 0.00646132 | L1 |
| rs1561090 | 11 | 30134621 | 1.84E-08 | 0.00739215 | L1 |
| rs17694317 | 11 | 30133539 | 1.85E-08 | 0.00620717 | L1 |
| rs10501124 | 11 | 30152823 | 1.85E-08 | 0.00646132 | L1 |
| rs10835627 | 11 | 30152440 | 1.86E-08 | 0.00646132 | L1 |
| rs10835628 | 11 | 30152472 | 1.86E-08 | 0.00646132 | L1 |
| rs1890826 | 11 | 30151369 | 1.87E-08 | 0.00646132 | L1 |
| rs12801198 | 11 | 30138657 | 1.87E-08 | 0.00620717 | L1 |
| rs11030974 | 11 | 30148027 | 1.88E-08 | 0.00624401 | L1 |
| rs11030962 | 11 | 30133383 | 1.88E-08 | 0.00599474 | L1 |
| rs12790814 | 11 | 30140647 | 1.88E-08 | 0.00620717 | L1 |
| rs12290734 | 11 | 30136951 | 1.89E-08 | 0.00599474 | L1 |
| rs12270725 | 11 | 30137281 | 1.89E-08 | 0.00599474 | L1 |
| rs11030968 | 11 | 30145566 | 1.92E-08 | 0.00624401 | L1 |
| rs1819675 | 11 | 30143725 | 1.92E-08 | 0.00624401 | L1 |
| rs10835621 | 11 | 30143185 | 1.93E-08 | 0.00624401 | L1 |
| rs2226272 | 11 | 30154694 | 1.93E-08 | 0.00646132 | L1 |
| rs10835630 | 11 | 30156918 | 1.93E-08 | 0.00646132 | L1 |
| rs7945546 | 11 | 30166958 | 1.94E-08 | 0.00624401 | L1 |
| rs3858477 | 11 | 30147651 | 1.94E-08 | 0.00624401 | L1 |
| rs7129554 | 11 | 30148451 | 1.94E-08 | 0.00624401 | L1 |
| rs4337008 | 11 | 30147324 | 1.95E-08 | 0.00603424 | L1 |
| rs12796334 | 11 | 30137835 | 1.97E-08 | 0.00578981 | L1 |
| rs7102959 | 11 | 30165497 | 1.98E-08 | 0.00603424 | L1 |
| rs12291589 | 11 | 30141437 | 2.02E-08 | 0.00603424 | L1 |
| rs10835622 | 11 | 30144850 | 2.04E-08 | 0.00583174 | L1 |
| rs12284112 | 11 | 30137361 | 2.06E-08 | 0.00599474 | L1 |
| rs10835620 | 11 | 30142421 | 2.07E-08 | 0.00583174 | L1 |
| rs6484476 | 11 | 30153418 | 2.08E-08 | 0.00583174 | L1 |
| rs17623478 | 11 | 30186296 | 2.08E-08 | 0.00632731 | L1 |
| rs1890825 | 11 | 30151032 | 2.10E-08 | 0.00575172 | L1 |
| rs11030998 | 11 | 30186969 | 2.14E-08 | 0.00607253 | L1 |
| rs12272116 | 11 | 30141466 | 2.15E-08 | 0.00563625 | L1 |
| rs713143 | 11 | 30161311 | 2.18E-08 | 0.00541652 | L1 |
| rs2884134 | 11 | 30154559 | 2.23E-08 | 0.00563625 | L1 |
| rs588929 | 11 | 30148605 | 2.33E-08 | 0.00526532 | L1 |
| rs12290971 | 11 | 30137360 | 2.33E-08 | 0.00595398 | L1 |
| rs930321 | 11 | 30144349 | 2.34E-08 | 0.00537063 | L1 |
| rs663173 | 11 | 30148309 | 2.37E-08 | 0.0050894 | L1 |
| rs11030961 | 11 | 30133062 | 2.37E-08 | 0.00486847 | L1 |
| rs475305 | 11 | 30144163 | 2.40E-08 | 0.0050894 | L1 |
| rs12800683 | 11 | 30129897 | 2.40E-08 | 0.00486847 | L1 |
| rs11030960 | 11 | 30132490 | 2.40E-08 | 0.00486847 | L1 |
| rs3847632 | 11 | 30139246 | 2.40E-08 | 0.00551252 | L1 |
| rs1865731 | 11 | 30131801 | 2.43E-08 | 0.00486847 | L1 |
| rs7935077 | 11 | 30131047 | 2.51E-08 | 0.00470317 | L1 |
| rs11030986 | 11 | 30163930 | 2.54E-08 | 0.0050894 | L1 |
| rs1369811 | 11 | 30130531 | 2.55E-08 | 0.00454365 | L1 |
| rs11030988 | 11 | 30164149 | 2.56E-08 | 0.0050894 | L1 |
| rs1361421 | 11 | 30164964 | 2.60E-08 | 0.00491955 | L1 |
| rs12282113 | 11 | 30166281 | 2.66E-08 | 0.00491955 | L1 |
| rs892952 | 11 | 30129570 | 2.66E-08 | 0.00438973 | L1 |
| rs12282163 | 11 | 30166358 | 2.68E-08 | 0.00475555 | L1 |
| rs11030985 | 11 | 30163848 | 2.71E-08 | 0.00475555 | L1 |
| rs12282076 | 11 | 30166411 | 2.72E-08 | 0.00475555 | L1 |
| rs1361424 | 11 | 30158843 | 2.74E-08 | 0.00475555 | L1 |
| rs11030987 | 11 | 30163939 | 2.89E-08 | 0.00444428 | L1 |
| rs10835624 | 11 | 30145081 | 2.94E-08 | 0.00468471 | L1 |
| rs1898174 | 11 | 30144693 | 2.96E-08 | 0.00468471 | L1 |
| rs1361422 | 11 | 30159138 | 3.09E-08 | 0.00429662 | L1 |
| rs11030982 | 11 | 30160231 | 3.09E-08 | 0.00429662 | L1 |
| rs3858478 | 11 | 30148707 | 3.13E-08 | 0.00437635 | L1 |
| rs11030970 | 11 | 30146288 | 3.18E-08 | 0.00437635 | L1 |
| rs17077610 | 13 | 83977056 | 1.04E-08 | 0.2361694 | L1 |
| rs1855622 | 13 | 83931406 | 3.73E-08 | 0.06657425 | L1 |
| rs4142268 | 13 | 83926866 | 4.01E-08 | 0.065088 | L1 |
| rs984749 | 13 | 83935236 | 4.23E-08 | 0.0636712 | L1 |
| rs983953 | 13 | 83930628 | 4.36E-08 | 0.05949754 | L1 |
| rs55756919 | 13 | 83946437 | 4.37E-08 | 0.06084377 | L1 |
| rs4409976 | 13 | 83940126 | 4.42E-08 | 0.06084377 | L1 |
| rs17077513 | 13 | 83936652 | 4.48E-08 | 0.05949754 | L1 |
| rs17077551 | 13 | 83943948 | 4.54E-08 | 0.05949754 | L1 |
| rs17077499 | 13 | 83931862 | 4.57E-08 | 0.05818415 | L1 |
| rs72628099 | 13 | 83953258 | 4.61E-08 | 0.05758611 | L1 |
| rs12429049 | 13 | 83922905 | 4.72E-08 | 0.05565255 | L1 |
| rs17077527 | 13 | 83939185 | 5.18E-08 | 0.05094838 | L1 |
| rs75347775 | 19 | 18385098 | 5.87E-49 | 0.18761969 | L3 |
| rs16982345 | 19 | 18389912 | 9.31E-48 | 0.67314559 | L3 |
| rs45543339 | 19 | 18392384 | 2.27E-47 | 0.1429242 | L3 |
| rs1227734 | 19 | 18390224 | 2.19E-08 | 0.4994183 | L1 |
| rs1054221 | 19 | 18389048 | 2.52E-08 | 0.37824243 | L1 |
| rs1227731 | 19 | 18387093 | 4.51E-08 | 0.125954 | L1 |
| rs12985909 | 19 | 18328573 | 1.29E-07 | 0.87860483 | L2 |
| rs7252185 | 19 | 18521415 | 4.50E-07 | 0.04925455 | L4 |
| rs8106332 | 19 | 18513669 | 4.83E-07 | 0.03783528 | L4 |
| rs6512271 | 19 | 18511889 | 4.97E-07 | 0.03693482 | L4 |
| rs10409408 | 19 | 18501085 | 5.05E-07 | 0.03693482 | L4 |
| rs57923655 | 19 | 18514341 | 5.05E-07 | 0.03605781 | L4 |
| rs10409346 | 19 | 18501011 | 5.06E-07 | 0.03605781 | L4 |
| rs12975407 | 19 | 18332840 | 5.07E-07 | 0.11515336 | L2 |
| rs4808808 | 19 | 18489372 | 5.08E-07 | 0.03693482 | L4 |
| rs8102717 | 19 | 18482916 | 5.31E-07 | 0.03520361 | L4 |
| rs8112372 | 19 | 18512322 | 5.33E-07 | 0.03236001 | L4 |
| rs7249760 | 19 | 18500735 | 5.34E-07 | 0.03520361 | L4 |
| rs4808812 | 19 | 18511633 | 5.55E-07 | 0.03356119 | L4 |
| rs8104247 | 19 | 18517395 | 5.56E-07 | 0.0343716 | L4 |
| rs1469413 | 19 | 18505714 | 5.68E-07 | 0.03277179 | L4 |
| rs4006662 | 19 | 18518302 | 5.78E-07 | 0.03356119 | L4 |
| rs10418331 | 19 | 18510860 | 5.83E-07 | 0.03277179 | L4 |
| rs4580317 | 19 | 18508927 | 5.95E-07 | 0.03200284 | L4 |
| rs7252130 | 19 | 18522626 | 6.32E-07 | 0.02913916 | L4 |
| rs8111397 | 19 | 18478235 | 8.06E-07 | 0.03515801 | L4 |
| rs11668704 | 19 | 18484825 | 8.45E-07 | 0.03352673 | L4 |
| rs4808807 | 19 | 18484026 | 8.49E-07 | 0.03352673 | L4 |
| rs11672385 | 19 | 18475949 | 7.22E-06 | 0.0136679 | L4 |
| rs10416993 | 19 | 18489645 | 8.87E-06 | 0.00830065 | L4 |
| rs7246788 | 19 | 18464640 | 8.93E-06 | 0.01570552 | L4 |
| rs7256530 | 19 | 18498280 | 9.00E-06 | 0.00830065 | L4 |
| rs4808805 | 19 | 18474600 | 1.01E-05 | 0.01085683 | L4 |
| rs11670392 | 19 | 18496970 | 1.03E-05 | 0.010276 | L4 |
| rs8112859 | 19 | 18512029 | 1.08E-05 | 0.00988122 | L4 |
| rs4808803 | 19 | 18473971 | 1.08E-05 | 0.01326301 | L4 |
| rs8106068 | 19 | 18514154 | 1.08E-05 | 0.00731313 | L4 |
| rs7247540 | 19 | 18473668 | 1.09E-05 | 0.01043368 | L4 |
| rs10410797 | 19 | 18504852 | 1.09E-05 | 0.00731313 | L4 |
| rs1469412 | 19 | 18505589 | 1.11E-05 | 0.00969106 | L4 |
| rs10423802 | 19 | 18457091 | 1.19E-05 | 0.00921811 | L4 |
| rs6512270 | 19 | 18507770 | 1.26E-05 | 0.00659133 | L4 |
| rs7507218 | 19 | 18525110 | 1.69E-05 | 0.00658279 | L4 |
| rs28534973 | 19 | 18424797 | 2.65E-05 | 0.01107733 | L4 |
| rs12162221 | 19 | 18524579 | 3.88E-05 | 0.02430217 | L4 |
| rs888669 | 19 | 18429834 | 4.47E-05 | 0.00767159 | L4 |
| rs172032 | 19 | 18522945 | 6.42E-05 | 0.01658845 | L4 |
| rs4808136 | 19 | 18508057 | 0.009762 | 0.00813991 | L4 |
| rs7256534 | 19 | 18475981 | 0.01019 | 0.00785985 | L4 |
| rs1806980 | 19 | 18455994 | 0.01056 | 0.00865641 | L4 |
| rs34256197 | 19 | 18495662 | 0.01162 | 0.00686144 | L4 |
| rs271622 | 19 | 18518467 | 0.01185 | 0.00678137 | L4 |
| rs8109573 | 19 | 18506341 | 0.0121 | 0.00662102 | L4 |
| rs271626 | 19 | 18516257 | 0.01219 | 0.00654524 | L4 |
| rs271623 | 19 | 18518137 | 0.01225 | 0.00654524 | L4 |
| rs76195 | 19 | 18519020 | 0.01227 | 0.00654524 | L4 |
| rs271621 | 19 | 18520522 | 0.01259 | 0.00676194 | L4 |
| rs7254249 | 19 | 18480935 | 0.0134 | 0.00750902 | L4 |
| rs11673604 | 19 | 18430178 | 0.01521 | 0.00903766 | L4 |
| rs8107743 | 19 | 18454229 | 0.01533 | 0.00634098 | L4 |
| rs10427083 | 19 | 18439466 | 0.01594 | 0.00628913 | L4 |
| rs1560119 | 19 | 18462865 | 0.01606 | 0.00627753 | L4 |
| rs8103660 | 19 | 18455585 | 0.01711 | 0.00852718 | L4 |
| rs10442 | 19 | 18434531 | 0.01768 | 0.00650762 | L4 |
| rs5994661 | 22 | 33050220 | 1.19E-08 | 0.4744308 | L1 |
| rs5754397 | 22 | 33049975 | 3.63E-08 | 0.14216557 | L1 |
| rs5754400 | 22 | 33061898 | 4.85E-08 | 0.10998865 | L1 |
| rs13057847 | 22 | 33095011 | 1.01E-07 | 0.06564407 | L1 |
| rs78618242 | 22 | 33098477 | 1.04E-07 | 0.06332465 | L1 |
| rs8135220 | 22 | 33097446 | 1.06E-07 | 0.06332465 | L1 |
| rs13057399 | 22 | 33094995 | 1.18E-07 | 0.05633954 | L1 |
| rs13090317 | 3 | 118584246 | 2.49E-08 | 0.27458076 | L1 |
| rs2970104 | 3 | 118531445 | 3.80E-08 | 0.01918 | L1 |
| rs4346538 | 3 | 118582268 | 5.52E-08 | 0.12880845 | L1 |
| rs6778385 | 3 | 118610381 | 6.75E-08 | 0.10348621 | L1 |
| rs6782220 | 3 | 118611504 | 7.04E-08 | 0.10032548 | L1 |
| rs6775687 | 3 | 118609930 | 8.97E-08 | 0.07277267 | L1 |
| rs1500078 | 3 | 118614948 | 1.25E-07 | 0.04271237 | L1 |
| rs9289112 | 3 | 118616799 | 1.26E-07 | 0.05301983 | L1 |
| rs6438468 | 3 | 118671728 | 1.37E-07 | 0.01768573 | L1 |
| rs7644018 | 3 | 118602586 | 2.80E-07 | 0.02914276 | L1 |
| rs4470512 | 3 | 118597872 | 2.81E-07 | 0.03272283 | L1 |
| rs9838729 | 3 | 118600096 | 3.81E-07 | 0.02161471 | L1 |
| rs9289110 | 3 | 118604427 | 3.94E-07 | 0.02245987 | L1 |
| rs9289111 | 3 | 118604688 | 4.30E-07 | 0.01957306 | L1 |
| rs9822639 | 3 | 118604069 | 4.68E-07 | 0.01936083 | L1 |
| rs60755482 | 3 | 119009385 | 0.0008174 | 0.05086338 | L2 |
| rs78491414 | 3 | 119007680 | 0.0008183 | 0.05086338 | L2 |
| rs16829222 | 3 | 118996122 | 0.0009789 | 0.05031001 | L2 |
| rs1016284 | 3 | 118987317 | 0.0009869 | 0.05411421 | L2 |
| rs1836689 | 3 | 118981233 | 0.0009899 | 0.05399843 | L2 |
| rs12053925 | 3 | 119016175 | 0.001017 | 0.04285272 | L2 |
| rs11922973 | 3 | 118981764 | 0.001035 | 0.05142664 | L2 |
| rs1435653 | 3 | 118986120 | 0.001044 | 0.05208396 | L2 |
| rs1836691 | 3 | 118979702 | 0.00106 | 0.05077879 | L2 |
| rs4687962 | 3 | 118975317 | 0.001068 | 0.05014026 | L2 |
| rs7636874 | 3 | 118968805 | 0.001079 | 0.05234197 | L2 |
| rs75572136 | 3 | 118957219 | 0.001281 | 0.04378727 | L2 |
| rs80015304 | 3 | 118980815 | 0.001283 | 0.04192052 | L2 |
| rs4687960 | 3 | 118937383 | 0.001384 | 0.04383789 | L2 |
| rs11715148 | 3 | 118934683 | 0.001397 | 0.04383789 | L2 |
| rs56232409 | 3 | 118926470 | 0.001494 | 0.03955357 | L2 |
| rs4687832 | 3 | 118911507 | 0.001655 | 0.03598619 | L2 |
| rs79221214 | 3 | 118914242 | 0.001725 | 0.03472072 | L2 |
| rs78317916 | 3 | 118898402 | 0.001848 | 0.03192823 | L2 |
| rs11709968 | 3 | 118924403 | 0.001881 | 0.03196843 | L2 |
| rs1879553 | 3 | 118896616 | 0.002083 | 0.02950238 | L2 |
| rs74887672 | 3 | 118901441 | 0.002173 | 0.02752234 | L2 |
| rs79151229 | 3 | 118925248 | 0.008198 | 0.00996638 | L2 |
| rs75535755 | 3 | 118971205 | 0.01455 | 0.0067102 | L2 |
| rs58781511 | 3 | 118996075 | 0.0183 | 0.00662888 | L2 |
| rs9861978 | 3 | 118967452 | 0.05238 | 0.00725144 | L2 |
| rs7613076 | 3 | 118980078 | 0.05788 | 0.00656832 | L2 |
| rs6786886 | 3 | 118963015 | 0.06011 | 0.00652837 | L2 |
| rs9312688 | 4 | 57486707 | 4.09E-22 | 0.2084262 | L1 |
| rs1441063 | 4 | 57485029 | 4.23E-22 | 0.19275259 | L1 |
| rs4865234 | 4 | 57488849 | 4.96E-22 | 0.17458361 | L1 |
| rs1441062 | 4 | 57483723 | 5.09E-22 | 0.11148704 | L1 |
| rs4865233 | 4 | 57488786 | 5.21E-22 | 0.17458361 | L1 |
| rs7683010 | 4 | 57492982 | 9.55E-22 | 0.06603618 | L1 |
| rs12210212 | 6 | 55337401 | 8.08E-09 | 0.14988321 | L2 |
| rs73436825 | 6 | 55331241 | 8.73E-09 | 0.1287361 | L2 |
| rs7768300 | 6 | 55335592 | 8.92E-09 | 0.13333216 | L2 |
| rs57378656 | 6 | 55340135 | 9.97E-09 | 0.11867165 | L2 |
| rs73436839 | 6 | 55338176 | 1.00E-08 | 0.11416617 | L2 |
| rs17683184 | 6 | 55333171 | 1.04E-08 | 0.10983867 | L2 |
| rs113706252 | 6 | 55341743 | 1.05E-08 | 0.10983867 | L2 |
| rs12209722 | 6 | 55340964 | 1.15E-08 | 0.09785377 | L2 |
| rs9370418 | 6 | 55401827 | 6.71E-06 | 0.19987526 | L1 |
| rs6900273 | 6 | 55400948 | 1.35E-05 | 0.08124715 | L1 |
| rs9382490 | 6 | 55405861 | 1.72E-05 | 0.06371642 | L1 |
| rs4715542 | 6 | 55404835 | 2.21E-05 | 0.03794343 | L1 |
| rs1546296 | 6 | 55401525 | 2.29E-05 | 0.03669871 | L1 |
| rs6912623 | 6 | 55406317 | 2.30E-05 | 0.03958912 | L1 |
| rs12201786 | 6 | 55403858 | 2.31E-05 | 0.03549861 | L1 |
| rs1472637 | 6 | 55402790 | 2.31E-05 | 0.03549861 | L1 |
| rs6920497 | 6 | 55400891 | 2.34E-05 | 0.03549861 | L1 |
| rs12205513 | 6 | 55405632 | 2.34E-05 | 0.03828542 | L1 |
| rs1502200 | 6 | 55400599 | 2.44E-05 | 0.03322576 | L1 |
| rs9349778 | 6 | 55402323 | 2.45E-05 | 0.03322576 | L1 |
| rs4715541 | 6 | 55398734 | 2.51E-05 | 0.03345904 | L1 |
| rs6918921 | 6 | 55400166 | 2.54E-05 | 0.03214992 | L1 |
| rs6913781 | 6 | 55399663 | 2.54E-05 | 0.03214992 | L1 |
| rs1502201 | 6 | 55400699 | 2.55E-05 | 0.03214992 | L1 |
| rs1502199 | 6 | 55400223 | 2.56E-05 | 0.03214992 | L1 |
| rs6936510 | 6 | 55399564 | 2.57E-05 | 0.03214992 | L1 |
| rs1472638 | 6 | 55402890 | 2.75E-05 | 0.02932596 | L1 |
| rs9367634 | 6 | 55400994 | 2.76E-05 | 0.03131698 | L1 |
| rs4715543 | 6 | 55405165 | 2.78E-05 | 0.03131698 | L1 |

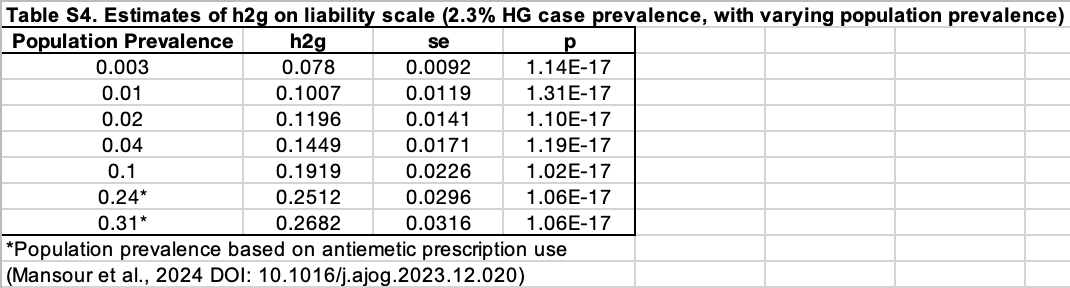

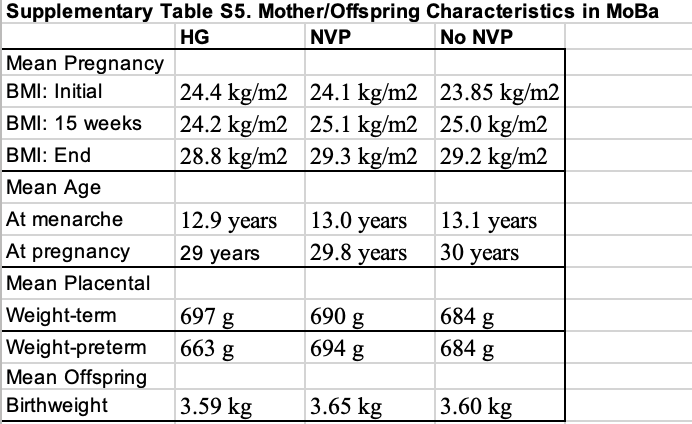

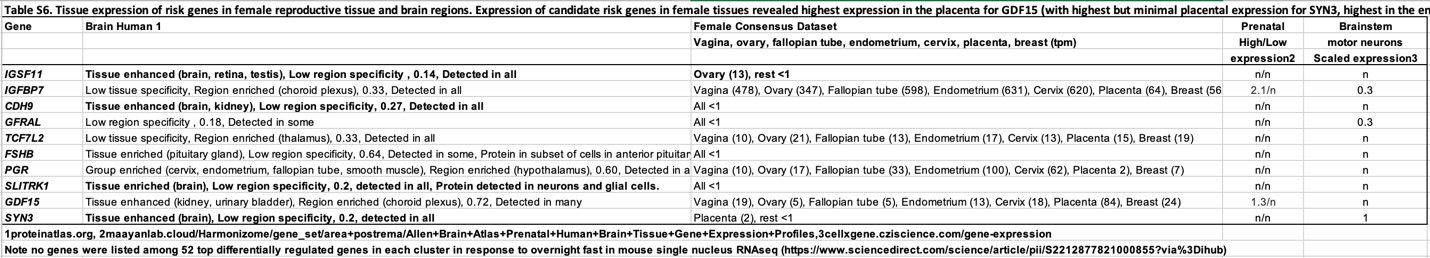
